# Dynamics of Eastern equine encephalitis virus during the 2019 outbreak in the Northeast United States

**DOI:** 10.1101/2023.03.06.23286851

**Authors:** Verity Hill, Robert T. Koch, Sean M. Bialosuknia, Kiet Ngo, Steven D. Zink, Cheri A. Koetzner, Joseph G. Maffei, Alan P. Dupuis, P. Bryon Backenson, JoAnne Oliver, Angela B. Bransfield, Michael J. Misencik, Tanya A. Petruff, John J. Shepard, Joshua L. Warren, Mandev S. Gill, Guy Baele, Chantal B.F. Vogels, Glen Gallagher, Paul Burns, Aaron Hentoff, Sandra Smole, Catherine Brown, Matthew Osborne, Laura D. Kramer, Philip M. Armstrong, Alexander T. Ciota, Nathan D. Grubaugh

## Abstract

Eastern equine encephalitis virus (EEEV) causes a rare but severe disease in horses and humans, and is maintained in an enzootic transmission cycle between songbirds and *Culiseta melanura* mosquitoes. In 2019, the largest EEEV outbreak in the United States for more than 50 years occurred, centered in the Northeast. To explore the dynamics of the outbreak, we sequenced 80 isolates of EEEV and combined them with existing genomic data. We found that, like previous years, cases were driven by frequent short-lived virus introductions into the Northeast from Florida. Once in the Northeast, we found that Massachusetts was important for regional spread. We found no evidence of any changes in viral, human, or bird factors which would explain the increase in cases in 2019. By using detailed mosquito surveillance data collected by Massachusetts and Connecticut, however, we found that the abundance of *Cs. melanura* was exceptionally high in 2019, as was the EEEV infection rate. We employed these mosquito data to build a negative binomial regression model and applied it to estimate early season risks of human or horse cases. We found that the month of first detection of EEEV in mosquito surveillance data and vector index (abundance multiplied by infection rate) were predictive of cases later in the season. We therefore highlight the importance of mosquito surveillance programs as an integral part of public health and disease control.

## Introduction

Eastern equine encephalitis virus (EEEV) is a mosquito-borne virus that causes periodic outbreaks in humans and horses in the United States (US) since its discovery in 1933 (Giltner & Shahan, 1933; TenBroeck & Merrill, 1933). The virus circulates in a bird-mosquito transmission cycle while infections of most mammals are considered “dead end hosts”. In humans, EEEV can cause severe disease, with an apparent case fatality rate of 30% and long-term neurological sequelae in more than half of those who survive (Lindsey et al., 2018). Still, diagnosed human cases are rare, with an average of 7 per year in the US. The largest human outbreak in more than 50 years was the 38 cases reported in 2019, including 12 deaths (Lindsey et al., 2020). The outbreak was not limited to the East Coast where cases are typically detected, as 10 of the human cases that year were reported from Michigan. The widespread EEEV cases in 2019 had significant impacts on the communities: many evening outdoor events were canceled to avoid mosquito exposure and aerial insecticide applications were the subject of public controversy (Shamus, 2019; Tunison, 2019). Thus, understanding the drivers of EEEV outbreaks and how to accurately communicate risk to the public is of high importance (Howard, 2019).

The key unanswered questions are (1) what factors facilitated the unprecedented EEEV activity in 2019, and (2) whether we can accurately estimate risk of human and horse infections? These are challenging questions to answer because the ecology of EEEV is complex, involving multiple species of bird and mosquito. *Culiseta melanura* serves as the main mosquito vector of EEEV in North America, lives in freshwater swamp habitats, and feeds primarily on passerine birds (Morris, 1988). Historically, *Coquillettidia perturbans* was implicated in the spillover process to humans as bridge vectors (i.e., vectors which feed on both birds and mammals) (Armstrong & Andreadis, 2022). This strict delineation between obligate avian and permissive feeders, however, is not so absolute, and *Cs. melanura* has also been found to occasionally feed on mammals in the Northeast (Molaei et al., 2006). In terms of viral dynamics, previous work demonstrated that EEEV circulates year round in Florida and is introduced into the Northeast through seasonal bird migration (Mundis et al., 2022; Tan et al., 2018), although this process does not happen predictably every year. Predicting the annual case dynamics is therefore difficult, having to take into account viral dynamics across multiple species and geographic scales.

In this study, we used a combination of phylodynamics, mosquito surveillance, and mathematical and statistical modeling to explore the dynamics of EEEV in the Northeast US, and specifically address factors behind the 2019 outbreak. We sequenced 80 EEEV isolates to add to the currently available genomic data, including 48 from 2019, and combined them with historical data to identify patterns influencing national and regional spread. We then explored which human, viral, mosquito, and ecological factors may contribute to years with many cases in humans and horses, with the aim of understanding if years like 2019 are predictable, or likely to be repeated. We confirmed that the 2019 outbreak was primarily driven by EEEV introductions from Florida rather than extended spread in the region. We also found that when there is regional spread in the Northeast it mostly originates from Massachusetts. We found no viral, human, or avian factors which contributed to the 2019 outbreak, but found that mosquito surveillance data was able to explain much of the variation in human and horse cases, highlighting the importance of high-quality and routinely collected mosquito data.

## Results

### High EEEV mosquito infections rates within known transmission foci

The outbreak of EEEV in humans and horses in 2019 was primarily focused in the Northeast US, defined as New York, Connecticut, Massachusetts, New Hampshire, Vermont, Rhode Island, and Maine. Using routine surveillance data from Massachusetts, Connecticut, and New York, we found that (1) cases occurred within previously known EEEV transmission foci and (2) the high number of human and horse cases in 2019 corresponded with a high number of trapped *Cs. melanura* and a high EEEV mosquito infection rate.

During 2019 there were 19 human and 26 horse cases reported in the Northeast US, compared to 11 human and 20 horse cases in 2005, the second largest outbreak since surveillance began in 2003 (**Fig. 1A**). The earliest human cases in the region were reported in July in Massachusetts. This is slightly earlier than the earliest Northeastern horse cases, which were reported in August across all Northeastern states other than Vermont (**Fig. 1B**). The last cases were in September in Connecticut and Massachusetts (human) and October in New York (horse).

**Figure 1.**
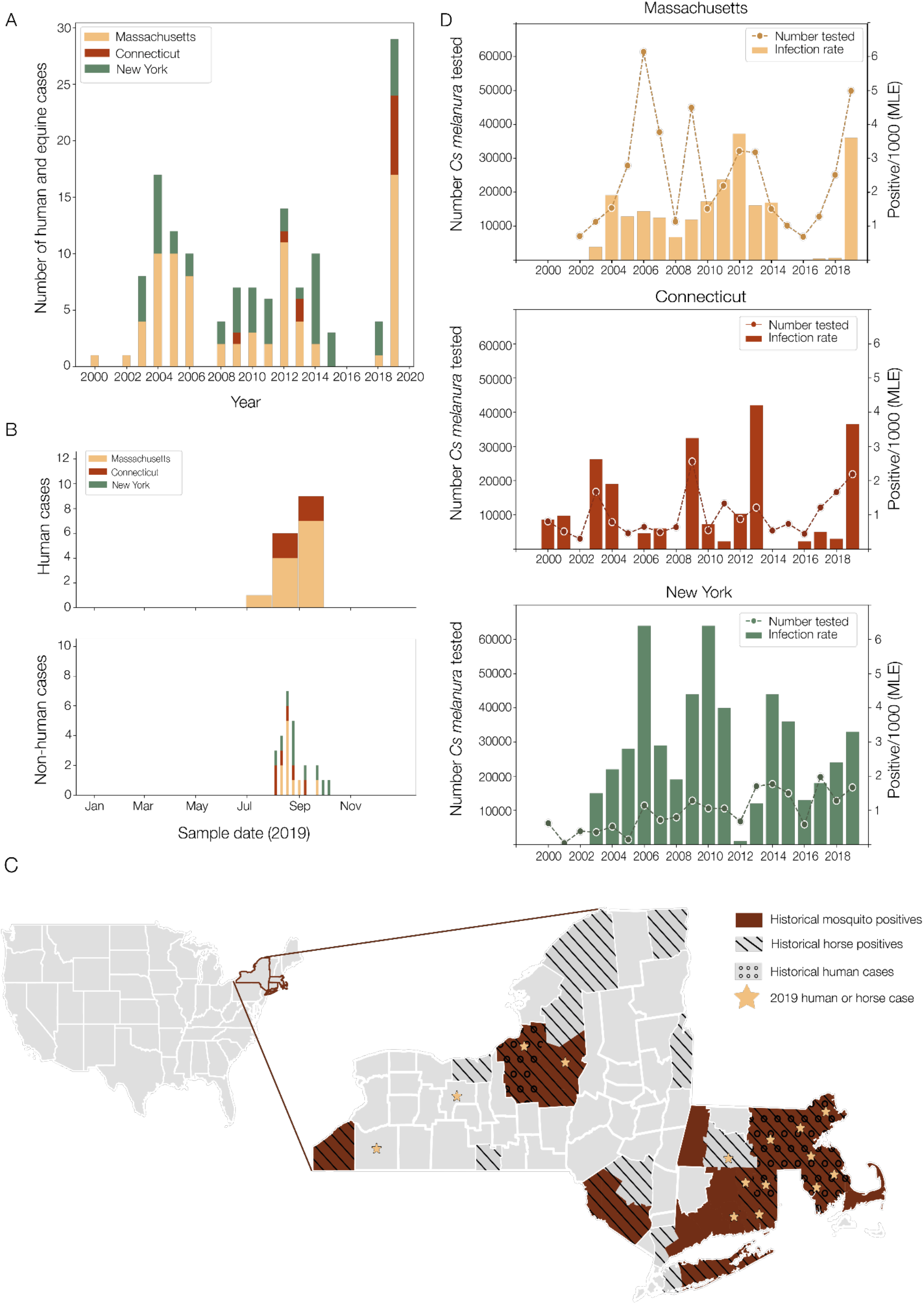
Temporal and geographical characteristics of previous EEEV outbreaks after 2000 and context of the 2019 outbreak in Massachusetts, Connecticut, and New York. A) Human and horse cases per year since 2000 in Massachusetts, Connecticut, and New York, colored by state. B) Human and veterinary cases in 2019 in these three states by sample date. Note that human cases are only available to the nearest month whereas veterinary cases have specific dates. C) Map showing geographical distribution of human and horse cases in 2019, shown as stars, relative to EEEV detections after 2000 in Massachusetts, Connecticut, and New York. Brown denotes counties where mosquito-positive pools have been sampled, hatched are non-human cases, and circles are human cases. D) EEEV infection rate of Culiseta melanura mosquitoes and the number tested in Massachusetts, Connecticut, and New York by year from 2000.

Historically, there have been two foci of transmission in the Northeast: an eastern, coastal focus which encompasses most of Massachusetts and Connecticut, and a western focus in central New York towards Lake Ontario. While cases were recorded in both foci in 2019, the vast majority were in the eastern focus in Massachusetts and Connecticut, with no human cases and only 8 horse cases reported in upstate New York (**Fig. 1C**). Cases primarily occurred in counties which had reported EEEV-positive human, horses, or mosquitoes before 2019. Further, the two counties which had not previously detected EEEV (Cattaraugus county in the southwest and Ontario county further north, both in central New York) are adjacent to counties which have (**Fig. 1C**). Therefore, there was not much geographical expansion in 2019, and cases fit into previously established transmission foci (**Fig. 1C**).

Using data from routine mosquito surveillance, we calculated the maximum-likelihood estimates (MLEs) of mosquito infection rates (see Methods). We found the number of tested *Cs. melanura* mosquitoes and EEEV infection rates in Massachusetts (MLE infection rate 2019 = 3.62/1000; 2003-2018 average = 1.21/1000, 95% CI: 0.67-1.75/1000) and Connecticut (MLE infection rate 2019 = 3.67/1000; 2003-2018 average = 0.94/1000, 95% CI: 0.36-1.52/1000), two states that had a high increase in cases compared to recent years (**Fig. 1A**), were high in 2019 (**Fig. 1D**). New York, which reported human and/or horse cases in 2014 (12 horse and 2 human cases), 2015 (3 human cases), and 2018 (3 horse cases), had more of a normal EEEV year in 2019 (8 horse cases). Subsequently, we did not find a high number of tested *Cs. melanura* mosquitoes and EEEV infection rates in New York in 2019 (MLE infection rate 2019 = 3.30/1000; 2003-2018 average = 2.49/1000, 95% CI: 1.53-3.45/1000). In general, the mosquito infection rate patterns in Massachusetts and Connecticut are more similar to each other compared to New York, which is likely indicative of the two distinct geographical foci of transmission described above.

### The 2019 outbreak in the Northeast was caused by several recent virus introductions from Florida

To explore the underlying spatial dynamics of EEEV in the US, in particular to determine the timing and the source of viruses causing the 2019 outbreak, we sequenced 80 isolates of EEEV from Massachusetts, Connecticut, and New York. Adding our newly sequenced EEEV genomes to the existing publicly available whole genome sequences provided a sufficiently detailed dataset to perform a phylogeographic reconstruction on a state level. We found that EEEV transmission in Florida routinely seeds other locations across the eastern seaboard and was the source of multiple lineages causing the 2019 outbreak.

Phylogeographic analysis to reconstruct virus spread requires sequence data from across spatial and temporal scales. Prior to the expansion of the nationwide arbovirus reporting system in 2003 (Lindsey et al., 2012), genomic sequence data for EEEV were relatively sparse. There are, however, some sequences from across the full range of years, with the earliest sequence from the first recorded outbreak in 1933 (**Fig. 2A**). Within the Northeast, EEEV sequencing was most concentrated on samples from southeastern Massachusetts and upstate New York (**Fig. 2B**). Nationally, most sequences were from the Northeast and Florida, although only until 2014 for the latter. There was sporadic sequencing as far west as Texas, although there are no sequences from many of the intermediate states along the east coast (**Fig. 2C**).

**Figure 2.**
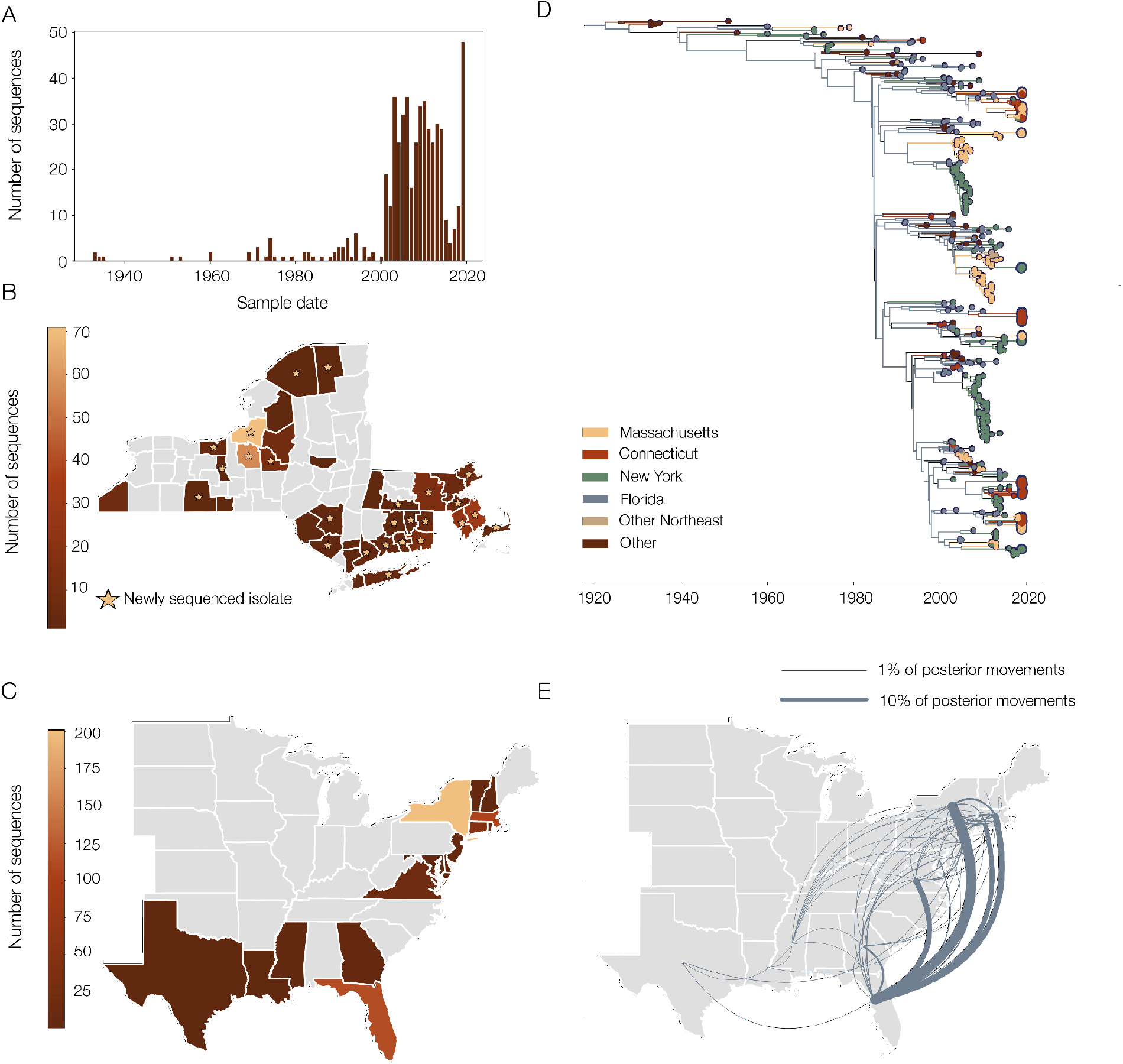
Phylogeographic reconstruction of EEEV from 2019 and prior outbreaks. A) Number of EEEV sequences in the dataset over time by year of sampling. Note that the nationwide reporting of surveillance data began in 2003. B) Location of EEEV sequences in the dataset from Massachusetts, New York, and Connecticut to the county level. Stars indicate the location of EEEV samples which were newly sequenced for this study. C) Location of all EEEV sequences in the study to state level. D) Time-resolved phylogeny colored by location of nodes from the discrete phylogeographic analysis. States in the Northeast are colored separately, but non-Northeast and non-Florida states are grouped together. Larger tips represent EEEV sequences from 2019. E) Movement of virus from the full posterior of the discrete phylogeographic analysis. Direction is anti-clockwise, and width of lines corresponds to frequency of movement across the posterior. Movements that make up fewer than 1% of the total posterior have been filtered out.

To provide additional geographical resolution within the Northeast and update the dataset to incorporate the 2019 outbreak, we sequenced an additional 80 isolates from across Massachusetts (n=17), Connecticut (n=38), and New York (n=25) (**Table S1**). They were primarily from *Cs. melanura* (n=63), although there were also isolates from other mosquito species: *Coquillettidia perturbans* (n=5), *Aedes vexans* (n=1), *Culex salinarius* (n=1), *Aedes canadensis* (n=1); as well as 9 sequences from horses and 1 from a turkey. These isolates were sampled from 2015-2019, with 48 from 2019, and were mostly from counties from which there were previously few or no sequences, particularly in Connecticut (**Fig. 2B**).

Combining our new EEEV sequences with publicly available data (531 total sequences), we performed a joint phylogeographic and phylogenetic reconstruction (**Fig. 2D**). We estimate the time of origin of the US phylogeny to be 1923.2 (95% Highest Posterior Density (HPD): 1920.5-1925.9), ten years before EEEV was first detected when it caused a large outbreak in horses in Virginia in 1933 (Giltner & Shahan, 1933). Further, we found a strong temporal signal for this dataset (**Fig. S1**) with an estimated evolutionary rate of 1.86×10^−4^ (95% HPD: 1.77×10^−4^-1.95×10^−4^) substitutions per site per year, in line with previous estimates (Tan et al., 2018).

Supporting previous results of phylogeographic analysis (Tan et al., 2018), we found that Florida forms the backbone of the phylogeny. In other words, EEEV transmission in Florida acts as a source of virus introductions into other states (**Fig. 2D**). When investigating the movement between states across the whole posterior distribution of the phylogeographic reconstruction, we found that 86.7% of EEEV movements start in Florida and end in every other state in the dataset (**Fig. 2E**). It is worth noting that there are not many sequences from states outside of Florida and the Northeast, and so movements involving states such as Texas, Georgia, and Virginia should be interpreted with caution, and there are likely more regional dynamics which we could not uncover with this dataset.

Using our new EEEV sequences, we found that the 2019 outbreak in the Northeast involved several independent virus introductions (**Fig. 2D**). Following the national trends, we infer that each of these introductions originated in Florida and were not related to other previous EEEV clusters sequenced from the Northeast. Therefore, the 2019 outbreak in the Northeast consisted of multiple EEEV lineages most likely introduced from the reservoir population in Florida, as opposed to long-term regional persistence or a single introduction with explosive growth.

### EEEV lineages do not typically persist longer than a few years in the Northeast

Having found that EEEV circulation in the Northeast, including the 2019 outbreak, is mostly driven by repeated introductions from Florida, we explored the maintenance of these lineages once they become established in the region. We found that viral lineages on average only persist for less than three years and are generally detected a year after introduction.

Due to the lack of sequences from Florida in general, but especially after 2014, the clades estimated by our discrete phylogeographic analysis are likely not precise estimates of the timings of EEEV introductions into the Northeast (**Fig. 2D**). In particular, we expect that we have estimated larger and longer-lived clusters after 2014 than there are in reality, as they have not been broken up by EEEV sequences from Florida. To account for this, we calculated the average branch length in the maximum clade credibility (MCC) tree in Northeastern clusters of more than three sequences with more than half of their sequences from 2014 or earlier, which was 1.28 years (standard deviation = 1.5 years). We then subdivided any clades that contained branches longer than the mean plus twice the standard deviation (4.3 years). This also included clusters from before 2014, indicating that sampling bias specifically involving Florida is an issue throughout the dataset. This procedure resulted in splitting the 61 EEEV introductions inferred from our discrete trait analysis into 75 separate introductions into the Northeast (**Fig. 3A**).

**Figure 3.**
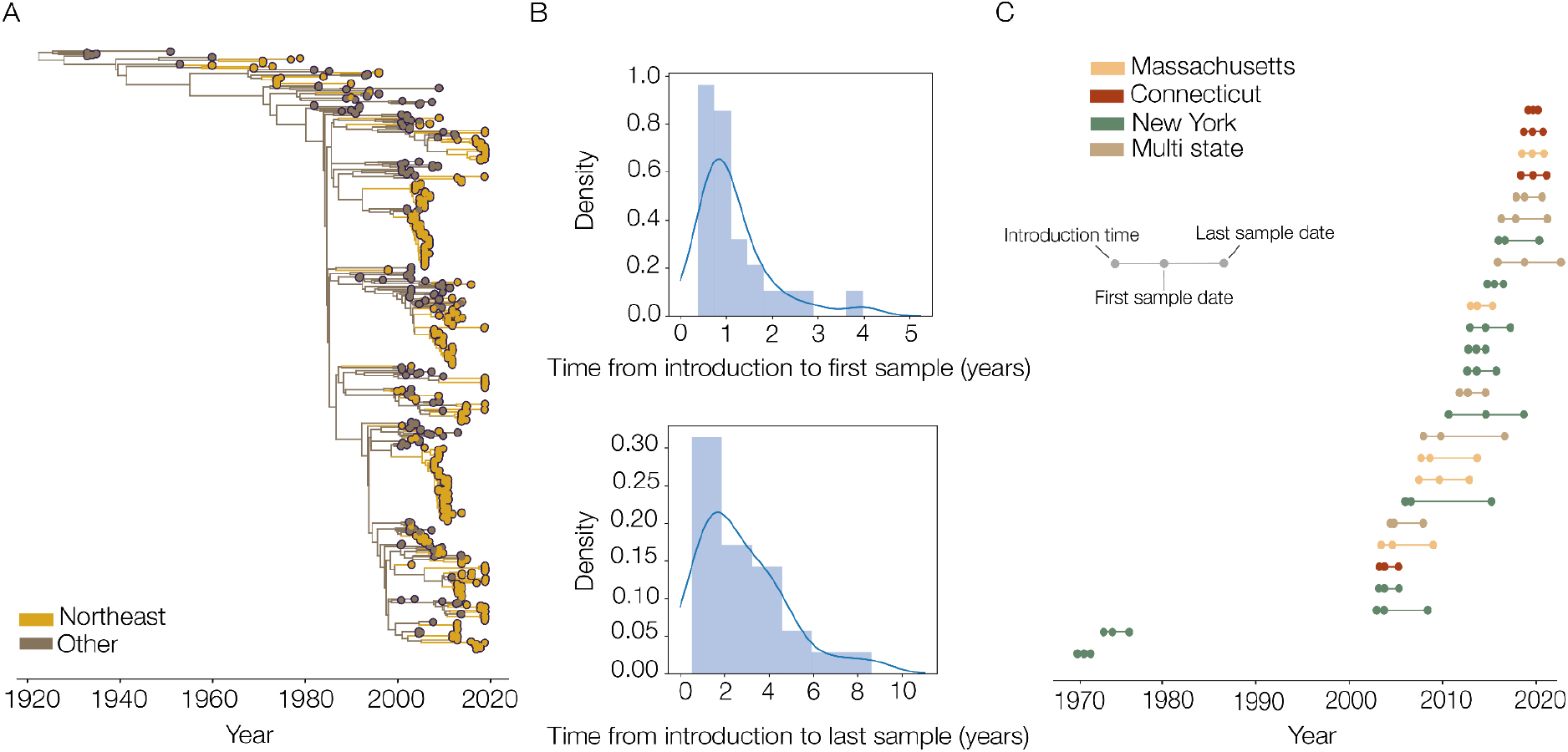
Detection and persistence of EEEV lineages in the Northeast. A) Time-scaled phylogeny estimating EEEV introductions into the Northeast, taking sporadic sampling into account by splitting up clusters with very long internal branches (see Methods). B) Distributions of time from introduction to first (top) and last sample (bottom) for Northeastern clusters of more than three EEEV sequences. C) Time of first node in the Northeast, first sample, and last sample for each cluster with more than three EEEV sequences, colored by state

Of these 75 introductions, we found 26 consisting of three or more sequences. On average, these took 1.17 (95% HPD: 0.46-3.27) years to be first detected in the genomic dataset, and circulated in the Northeast for an average of 2.8 (95% HPD: 0.65-7.5) years after being introduced (**Fig. 3B**). This supports previous analysis of the region showing limited multi-year maintenance in the region (Oliver et al., 2020; Young et al., 2008). The groups of lineages can be seen to roughly follow three wave patterns in the 2000s, between 2010 and 2014, and then 2014 to 2019 (**Fig. 3C**), possibly corresponding to infection and immunity patterns within the bird population in the region.

Of the 26 larger Northeastern introductions, 8 included samples from the 2019 outbreak, and half of these were solely composed of 2019 samples (**Fig. 3C**). We estimate that the average introduction time of these 2019 clusters was in mid 2017 (95% HPD: 2015-11-14 to 2019-01-15), and the earliest introduction was late 2015. Therefore, the EEEV lineages sequenced from 2019 were introduced no more than 4 years before the outbreak and after the most recent Northeast EEEV outbreak in 2014.

### Multi-state spread of EEEV originating in Massachusetts

Considering that EEEV lineages have a short lifespan in the Northeast (**Fig. 3**), we subsequently investigated the extent to which they could spread within the region during that time. We found that most of the Northeast EEEV clusters were detected within a single state, suggesting that inter-state regional spread is rare. Where we did detect between-state spread, however, Massachusetts appeared to be an important regional source.

Of the 26 Northeast clusters with three or more EEEV sequences (**Fig. 3C**), most (n=19) were from a single state, and only a single cluster was found in three states despite the density of sequencing in the region. Of the 7 clusters with sequences from multiple states, we found that most viral movements were inferred to be from Massachusetts to other states (7 out of 8 movements in the MCC tree, **Fig. 4A**) and the origin of 5 out of 6 subtrees was in Massachusetts (**Fig. 4B**). The subtree that does not have an origin in Massachusetts circulated mostly in Connecticut but does contain a sequence from Massachusetts. Importantly, multi-state clusters were not necessarily sampled in neighboring counties (**Fig. 4B**), indicating true spread within the region, possibly by infected birds.

**Figure 4.**
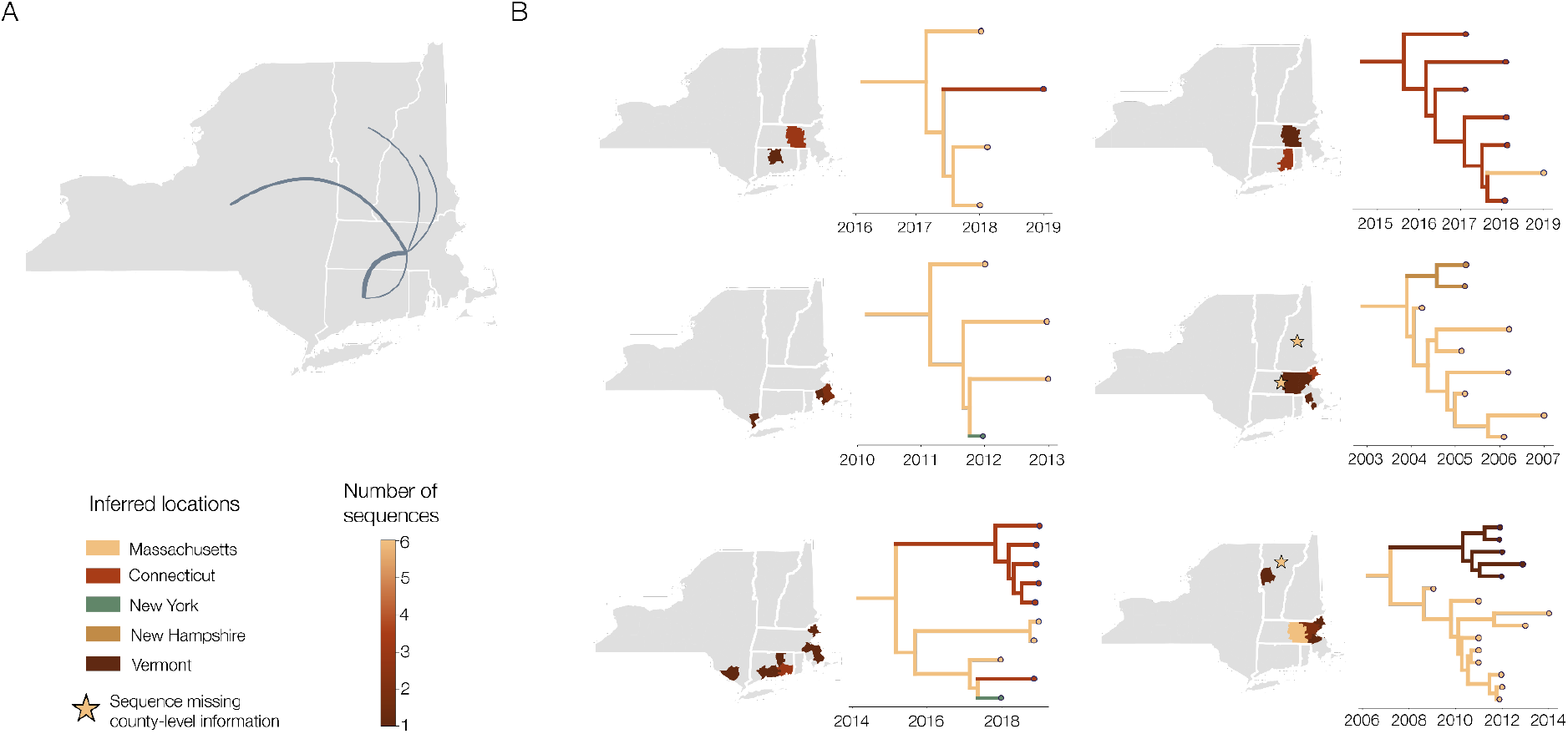
Phylogeographic spread of EEEV within the Northeast. A) Between-state movements from the maximum clade credibility (MCC) tree of the phylogeographic analysis within the six Northeastern EEEV clusters which have between-state movement. Width of lines relates to the number of movements, and direction is anti-clockwise. Maps showing sampling location to the county level for each EEEV cluster with multiple states where available. Stars indicate where there are sequences with information only at the state level (n=2 in New Hampshire, n=1 in Massachusetts and n=3 in Vermont). Corresponding subtrees are shown to the right of each map, colored by inferred location to the state level.

It is worth noting that only two of these multi-state clusters contain sequences from New York, despite the majority of EEEV sequences being from that state, and its size and central location in the region. These New York sequences were from the southeast part of the state (**Fig. 4B**), closer to the prominent Massachusetts-Connecticut focus rather than the transmission clusters in upstate New York. Therefore, while limited spread is happening in the region, it primarily only involves the eastern part of the region where smaller states are close together.

While none of the multi-state clusters were solely sampled in 2019, we found that half of them contain EEEV sequences sampled in 2019 (**Fig. 4B**). Therefore, the 2019 outbreak appears to have a wider within-cluster geographical spread than previous ones (e.g., 2012-2014 sequences are only found in 2 subtrees; **Fig. 4B**). The spread in 2019 was possibly driven by the high numbers of infected mosquitoes (**Fig. 1D**) leading to more infected birds and more opportunities for cross-state movements. We emphasize, however, that cases were mostly reported in counties that had previous EEEV cases (**Fig. 1C**), and so this was strictly a within-cluster effect and the outbreak as a whole does not show geographical expansion.

### Associations of virus, human, and bird factors with EEEV outbreaks were not found

The general epidemiology underlying the 2019 EEEV outbreak appears to be similar to previous outbreaks, with short-lived introductions from Florida driving transmission in the Northeast (**Fig. 2-3**), and infections occurring in similar locations (**Fig. 1**). Therefore the change in the lead-up to the 2019 outbreak, which underlies the increase in cases, may be due to mosquito population, virus evolution, human behavior, or bird populations, and there are multiple possible factors that are not mutually exclusive. Determining which factors were important for the 2019 outbreak may help us to understand what drives EEEV outbreaks in general. First, we ruled out that the 2019 outbreak was driven by virus evolution. Then, we examined case demographics and aspects of human behavior, and we did not find any associations between the age, sex, or timing of human cases to outbreak years, nor were humans spending more time outside in 2019 compared to 2018. Finally, we did not find strong evidence for the population sizes of key bird species as a correlate of cases or mosquito infection rates, though we are lacking important data on bird immunity to EEEV. Together, with the data available to us, virus genomics, case demographics, and bird population sizes fail to explain or predict EEEV outbreaks.

#### Intrinsic viral factors and effective population size

A major concern when any pathogen leads to a large increase in case counts is that it has evolved to have a higher virulence or transmissibility. In the case of EEEV, increased transmissibility would refer to a higher infection success in either mosquitoes or birds. Increased virulence would lead to increased case ascertainment, as diagnosis occurs on hospitalized individuals and most human infections are probably asymptomatic (Morens et al., 2019). We would expect to see a phylogenetic signal for any intrinsic variability in transmissibility or virulence.

We searched the alignment for any nucleotide substitutions, relative to a reference sequence from 2005, which are common to some or all of the 2019 EEEV sequences. There were 17 substitutions across the genome shared by all of the 2019 sequences, a further two that more than 90% of them shared, and an additional two that 75% of them shared. These 21 substitutions, however, were found across the whole phylogeny, in at least 93% of non-2019 EEEV sequences. We found no shared substitutions unique to more than 75% the sequences sampled in 2019. This was expected given that these sequences are spread across the phylogeny (**Fig. 2D**), as there would have to be extremely strong positive selection leading to multiple instances of convergent evolution for unique shared substitutions to be possible.

Past population size of viruses can be useful in inferring underlying transmission dynamics. In the case of EEEV, this can be done using a skygrid model (Gill et al., 2012), which we used to estimate changes in virus effective population size (in birds and mosquitoes) over time across the whole of the US. We found that there was a steady increase in effective population size in the 1990s, which then plateaued and decreased more recently in the late 2010s (**Fig. S2A**).

To explore if this could be formally connected to EEEV infections in humans and horses, we used an extension of the skygrid model to incorporate case data as covariates (Gill et al., 2016). We hypothesized that increased transmission in the mosquito-bird cycle could lead to more infections and reported cases in humans and horses. We tested 9 different covariates independently using data from 2003 onwards: all human and horse cases, only horse cases and only human cases each with no lag, a 1 year lag or a 2 year lag (**Fig. S2B**). We did not find any significant associations between the viral population size and any of these covariates. There is, therefore, no simple association between national circulation and reported cases, meaning that regional dynamics are likely more important for regional cases. More recent sequencing data in general, particularly from the Northeast to allow the inference of population dynamics around the outbreak in 2019, could confirm this.

#### Case demographics and human behavior

Human behavior could be one explanation for the increase in human EEEV cases in 2019, which could be revealed by analyzing the case demographics. Since 2003, we found that the cases have been predominantly male (including in 2019, where 62% of the cases were male), and this proportion was not found to vary across years (Fisher’s exact test, p=0.838, **Fig. S3A**). In comparison, the proportion of cases in different age categories were found to vary across years (p=0.047, **Fig. S3B**). To determine if this variation was meaningful, we divided years with cases since 2003 into large outbreak years (5 or more human cases) or small outbreak years. When we compared these two groups, we did not find any evidence of a difference in age distribution (Fisher’s exact test, p=0.322). Finally, we compared the timing of human and horse cases in the Northeast. The median month of symptom onset in all years was either August or September, with earliest cases in July and latest in October. We tested whether the median month of infection correlates with the number of cases and found that it does not (Pearson’s correlation, p=0.777, **Fig. S3C-D**).

To further explore whether human behavior may have changed in 2019, leading to a higher EEEV exposure rate, we compared the length of time individuals spent outside in relevant counties in Massachusetts, Connecticut, and New York (**Fig S3E**). We performed a paired t-test on the indoor activity seasonality metric for each of these counties in June, July, August, and September between 2018 and 2019 (data only available for these years) using data from (Susswein et al., 2022). We found no evidence of a difference in indoor-ness, and by extension the reciprocal outdoor-ness, between 2018 and 2019 in counties that had ever had a human case of EEEV (p=0.56) or only those which had a case in 2019 (p=0.72). Therefore, human behavior, the timing of cases, and case demographics do not appear to be the primary drivers of EEEV outbreaks based on our analysis of the available data. Although, understanding the behaviors and activities linked to human exposure is still an important area of research.

#### Bird populations

Bird populations likely have a complicated role in EEEV outbreaks. While the presence of competent bird species is a necessity for EEEV transmission, abnormally large bird populations may dilute the virus, decreasing the likelihood of mosquito infection during blood feeding (Kramer & Ciota, 2015). Moreover, when sufficient opportunities exist for mosquitoes to feed on birds, it may limit the pursuit of mosquitoes to feed on alternate sources, such as humans and horses. Finally, bird population immunity is likely a key component of the potential for EEEV perpetuation, but studies that routinely collect bird serology data are difficult and not often conducted.

While data on birds are limited, we examined the effect of the (1) absolute abundance and (2) proportion relative to all species of 8 bird species for which *Cs. melanura* has a high blood feeding preference (Armstrong & Andreadis, 2022; Molaei et al., 2016) on (A) human and horse cases and (B) mosquito EEEV infection rate using a Poisson regression. We also compared these metrics for 8 bird species for which a low feeding preference was estimated (Molaei et al., 2016) as a control (**Fig. S4**). In New York, we found very few significant relationships between the abundance or proportion of any of the high-preference bird species to case counts or infection rates. There are, however, several significant relationships in the low-preference bird species, suggesting that this comparison is not valid for New York. Further, the comparisons to mosquito infection rate almost all have high p-values regardless of state, bird species, or metric. In Massachusetts and Connecticut, we found several positive relationships between high-preference bird species and case counts, notably the common grackle (abundance only; p<0.001 for both states), chipping sparrow (proportion only; p<0.01 for both states), tufted titmouse (proportion only; p<0.01 for both states), and warbling vireo (abundance and proportion; p<0.01 for both states). There are, however, some low-preference birds with significant relationships (although fewer), meaning that the relationship between specific bird presence and EEEV transmission/cases is complicated and not easily solved here.

### EEEV outbreaks are primarily driven by the infection dynamics within mosquito populations

After ruling out or not finding associations with other factors, we subsequently focused on mosquitoes where we have detailed temporal data. After the CDC expanded the arboviral surveillance reporting system (ArboNET) and federal funding to the states in the early 2000’s (Hadler et al., 2015), many states began to routinely trap and test mosquitoes for multiple viruses, including EEEV and West Nile virus. This provides a glimpse into the transmission cycle of the virus, and allows us to explore if mosquito populations or infection rates are associated with outbreaks. We used mosquito surveillance data from all three states, and detailed data from Connecticut and Massachusetts, to develop a transmission model. Overall, we found that a high abundance of *Cs. melanura*, connected to environmental factors, is necessary, but not sufficient, for an outbreak of EEEV in humans and horses. When infection rates are included, however, these two metrics together are predictive of overall human and horse cases. We then applied this model to obtain predictions of total human and horse cases based on early season measurements.

To begin with, we found when there were more EEEV-positive *Cs. melanura* pools (i.e. mosquitoes were tested in groups of up to 60 from the same trap and date), there were more human and horse cases in Massachusetts, Connecticut, and New York. (**Fig. 5A**). Further, in 2019, there were more positive mosquito pools detected in Massachusetts and Connecticut than previously recorded. In comparison, the number of EEEV-positive mosquito pools detected in New York was more within the expected range of historic values, which aligns with the normal number of case counts in 2019 for that state. It must be noted that Massachusetts and Connecticut practice reactive sampling, and so more mosquito traps are set when EEEV-positive mosquitoes are found. To account for this, we also found the same strong correlation between the *Cs. melanura* EEEV infection rates (MLE per 1000 mosquitoes, see methods) and case counts (**Fig. S5A-B**). Therefore, our data suggest that human and horse cases are primarily driven by the abundance of infected mosquitoes more so than behaviors that increase exposure to mosquitoes.

**Figure 5.**
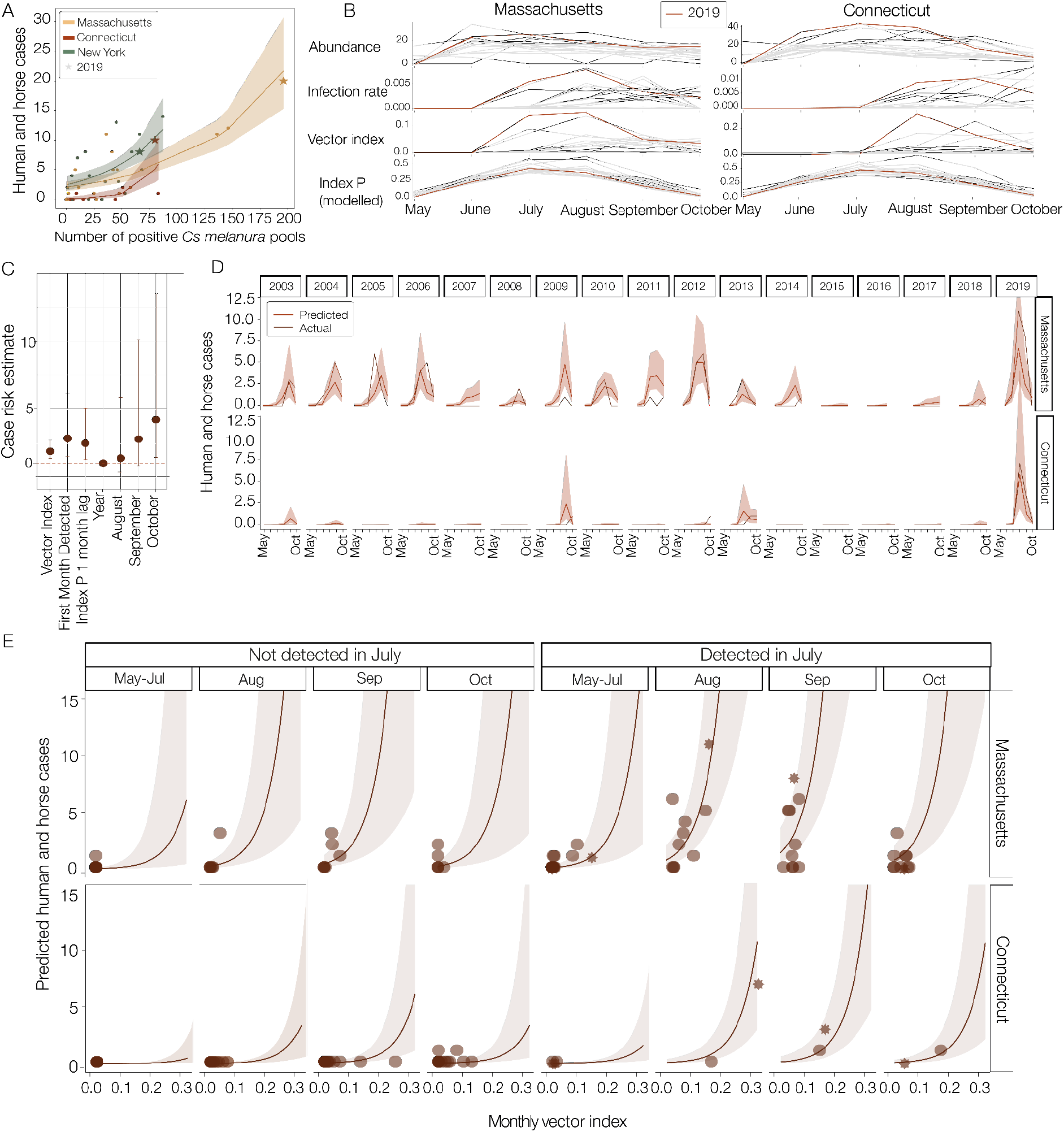
Model predictions of EEEV cases using mosquito infection estimates. A) Poisson regression by state of the number of EEEV positive Cs. melanura pools sampled and human and horse cases. B) Monthly trends across the year of mosquito abundance (Cs. melanura collected per trap per day), infection rate (MLE per 1000 Cs. melanura), vector index (abundance multiplied by the infection rate) and index P (modeled estimate of reproduction number), for Connecticut and Massachusetts. 2019 values are highlighted in red. C) Results of a negative binomial regression model for both Connecticut and Massachusetts combined of case risk predicted by vector index, first month of EEEV detection, index P with a 1 month lag, year, and risk compared to the month of May-July of cases in August, September, and October. The covariate for state was removed for scale, as its estimate is 11.95 (95% CI: 3.88 to 43.70) with Connecticut being the reference group. D) Actual and predicted case counts for each year based on the modeled results presented in C. Shaded areas denote error in the predicted values. E) Simulations using the previously described model. Predicted cases and intervals utilized a range of plausible vector index values and whether EEEV was detected in July or not, with index P held constant using the average value derived from the years studied for each month; and split by state. Shaded areas represent 95% confidence intervals and points are real data points from the EEEV mosquito surveillance program discussed elsewhere. Stars indicate values from 2019 and darker points indicate the actual modeled value of Index P.

To further explore the within-season EEEV dynamics, we focused on Connecticut and Massachusetts, for which we have more detailed mosquito data. We plotted the monthly *Cs. melanura* abundance (mosquitoes/trap/day), EEEV infection rate, and vector index (abundance multiplied by the infection rate to estimate the relative number of infected mosquitoes (Fauver et al., 2016; Nasci et al., n.d.) for each state, highlighting 2019 (**Fig. 5B**). While there were considerable yearly variations in both states, in general *Cs. melanura* abundance rapidly increased in early summer and tended to peak by June to July. The EEEV infection rates lagged behind, peaking in August or September, with infected mosquitoes often being detected in Massachusetts earlier than Connecticut. Vector index, therefore, tended to rise earlier (July) and persist longer in Massachusetts than Connecticut, though it often peaks in August in both states. In 2019, we found that both the *Cs. melanura* abundance and EEEV infection rates were high in both states, leading to the highest vector index values in the 17-year dataset (**Fig. 5B**). Thus the 2019 EEEV outbreak was driven by environmental conditions that supported larger *Cs. melanura* populations and transmission dynamics that led to high infection rates.

We next investigated whether environmental data could help explain the high abundances or EEEV infection rates. To explore this, we used index P (Obolski et al., 2019), a modeled estimate of reproduction number of mosquito-borne viruses based on climate factors, including temperature and humidity, by including prior estimates for *Cs. melanura*-borne transmission of EEEV (**Table S2**). Our assumption was that index P would best model the vector index. While our index P estimates usually peaked in July to August, similar to the vector index, we found that it actually correlated most closely with the *Cs. melanura* abundance dynamics (**Fig. S5C**; correlation coefficient of 0.57, p-value < 0.0001 for Connecticut, 0.24 and p<0.05 for Massachusetts). Furthermore, our modeled index P in 2019 was relatively average across the 17 years in both states (rank 9/17 for Connecticut, 7/17 for Massachusetts). Thus, for EEEV in the Northeast, index P is useful for describing the seasonal dynamics of transmission, but the temperature and humidity variables alone are not capable of describing which years will have a high number of EEEV-infected mosquitoes.

To interrogate further how different factors combine to explain cases, we built a negative binomial regression model that uses index P, vector index, state, month of first EEEV detection, the year, and the month to explore counts of total human and horse cases (**Fig. 5C-D, Table S3**; results from different variations of the model are shown in **Fig. S6**). Because of the yearly variation in EEEV cases (**Fig. 5D**, see “Actual” cases), we found that year and being in the month of August or September lead to no significant changes in risk of cases (**Fig 5C**). Vector index (2.07, 95% CI: 1.5-3.0), the month of first detection compared to June (3.17, 95% CI: 1.34-7.92), index P with a 1 month lag (2.31, 95% CI: 1.19-4.65), and being in the month of October (4.18, 95% CI: 1.39-13.55), however, all lead to a significant increase in the risk of human and horse cases (**Fig. 5C**). Further, being in Massachusetts as compared to Connecticut had the largest increase in risk (11.95, 95% CI: 3.88-43.70). When used to fit cases each year, our modeled estimates tracked closely with observed cases, where cases with the model being off on average by 0.47 cases per observation (Mean Absolute Error (MAE) = 0.47) and had strong explanatory power (Nagelkerke R^2^ = 0.80; **Fig. 5D**). Importantly, vector index was an important driver of the outbreak in 2019, where elevated levels correspond well with high case counts.

Next, we applied this model to examine if there were early season predictors which might indicate an increased risk of horse and human cases later in the season (**Fig. 5E**). While index P varies from year to year and is connected to mosquito abundance, it does not correlate well with vector index as it cannot predict when EEEV is present in the Northeast (**Fig. S5C**). We therefore held index P constant and simulated vector index across a range of values to predict the total human and horse cases for the year, and to explore its relationship with the month of first detection. We found that our model underpredicts cases when the vector index is low, as the real data points fall outside of the prediction intervals (**Fig. 5E**), indicating a potential lag in identification of EEEV cases. This could be due in part to stochastic chance in surveillance programs missing EEEV with low levels of virus circulation. As the vector index increases, however, our predicted values become closer to the observed data points.

The timing of the first detection of EEEV in mosquitoes is also important, and of particular note is that years with high case counts always first detected EEEV in July (**Fig. 5E**). This makes intuitive sense as the virus has more time to circulate to high levels in birds while there are sufficient mosquitoes present for it to transmit to humans. While the interaction between first EEEV detection in July and the vector index is not significant, this could be in part due to no cases being detected in years when the vector index was high but EEEV was first detected after July. The impact of the timing of the vector index is clear however, with a 220% increase in risk of cases when EEEV is detected in July (**Fig. 5E**). This is reflected in the empirical data, as the maximum number of cases in years when EEEV is detected after July is only 5.

Our negative-binomial regression model could be used as a tool for mosquito surveillance programs to estimate the risk of cases, which typically are reported in August-October, based on the timing of first detection and the vector index. For example, if EEEV is detected in mosquitoes in July in Massachusetts, and the vector index in August is 0.15 (similar to the value measured in 2019), we predict 7.0 human and horse cases in total that year (95% CI: 2.9-16.6), compared to 2.2 cases (95% CI: 0.6-7.9) if EEEV is detected after July during a typical summer season. In Connecticut, the same situation would correspond to an expected 0.6 cases (95% CI: 0.2-1.8) for July detection and 0.2 cases (95% CI: 0.1 to 0.6) for a later detection. With a more extreme August vector index of 0.30 in Massachusetts (higher than has ever been measured), our model predicts 89.2 cases (95% CI: 17.0-468.5) if EEEV is detected in July versus 28.2 cases (95% CI: 3.4-231.3) cases being detected later. While these predictions have wide confidence intervals, they provide a relative estimate of the possible extent of the cases in a given year, and therefore an indication of how much effort should be put into various mosquito control measures.

Finally, while all of our previous results focused on *Cs. melanura, Coquillettidia perturbans* is also thought to be an important bridge vector for infections in horses and humans (Armstrong & Andreadis, 2022; Sherwood et al., 2020). This species is more abundant on average in Connecticut than *Cs. melanura* (**Fig. S5D**), with populations peaking in late June. However, only 9 *Cq. perturbans* pools were positive for EEEV from 2003-2019 in the state, or 1.9% of all positive pools; and it was only the 8th most commonly positive mosquito species for EEEV. In 2019, there were six EEEV positive pools of *Cq. perturbans* sampled in Connecticut, making it the third most commonly sampled positive mosquito species for the year, although it still only made up 5% of the positive mosquito pools. This does vary between states, however, as *Cq. perturbans* made up a third of positive pools in Massachusetts in 2019 (Armstrong & Andreadis, 2022). Therefore, *Cq. perturbans* may still have a role as a bridge vector, but the transmission dynamics of EEEV in these states is primarily driven by *Cs. melanura*.

## Discussion

The 2019 EEEV outbreak in the Northeast US was exceptional in that it had the highest number of human cases recorded in more than 50 years, along with dense *Cs. melanura* vector populations with high virus infection rates (**Fig. 1 & 5**). We found that other components of 2019 were similar to outbreaks of years past. We showed that EEEV is not persistently maintained in the Northeast, with virus lineages typically going extinct within the region within a few years (**Fig. 3**). Instead, the virus is frequently reintroduced into the region from Florida where EEEV is endemic (**Fig. 2**). Thus, EEEV outbreaks in the Northeast are limited by stochastic and deterministic factors that support new virus introductions, and to habitats that are suitable for the mosquito vectors. We identified two primary transmission foci in the Northeast: (*1*) an eastern, coastal focus which encompasses most of Massachusetts and Connecticut, and (*2*) a western focus in central New York towards Lake Ontario (**Fig. 1**). The 2019 outbreak primarily occurred in the former, especially in regards to human cases. We found that human and horse cases are associated with a high vector index (large number of EEEV infected *Cs. melanura* mosquitoes), and we constructed a model using environmental and mosquito surveillance data that could estimate cases (**Fig. 5**). Finally, we found that a high early season vector index can be used by surveillance systems to predict human EEEV risk and direct control efforts (**Fig. 5E**). It remains unclear as to what causes high EEEV infection rates in mosquitoes, and therefore what contributed to the exceptionally high rate in 2019.

We conducted a phylodynamic analysis of our 80 newly-generated EEEV genome sequences combined with historical samples to explore the dynamics of EEEV in the Northeast of the US, with a focus on 2019 (**Fig. 2**). As both EEEV isolates and extracted RNA are designated as Select Agents in the US, it is extremely difficult to obtain clearance to store, transport, and in this case, sequence them. Thus our genomic dataset is critical to support future EEEV research. Using these additional sequences, we confirmed earlier studies which identified Florida as the source of EEEV introductions into the Northeast and other parts of the US (Tan et al., 2018) (**Fig. 2**). The documented endemic transmission in Florida and the limited number of EEEV cases reported outside of the US add further support to the hypothesis of Florida as a source for the Northeast. EEEV sequences from outside of the US, however, are limited in number (n=3) and we did not include these data in our analysis. Therefore, we cannot rule out a non-US external source of introductions into the Northeast. Further sequencing from other countries, enabling a broader, regional analysis of EEEV would allow the investigation of the international dynamics of EEEV and the identification of at-risk regions.

We found that while interstate spread of EEEV within the Northeast is not common, Massachusetts appears to be an important focus for when it does happen (**Fig. 4**). This may be in part due to bird migration routes, more consistent EEEV activity in Massachusetts compared to the surrounding states (**Fig. 1 & 5**), and/or that EEEV infection in mosquitoes in Massachusetts increases earlier in the year than it does in Connecticut (**Fig. 5**). Thus, controlling EEEV transmission in Massachusetts may help to alleviate some EEEV transmission in other states.

In discussing the movement of EEEV across the US, it is important to note that there is significant sampling bias in our dataset, in terms of both time and location of the sequences. Most of the sequences are from the Northeast US, and there are no sequences from Florida after 2014. While we have attempted to mitigate the latter point by splitting Northeastern clusters with long branches (see Methods), geographical heterogeneity can lead to overconfidence in the transition times (Layan et al., 2023). Further, the lack of EEEV sequences from other East Coast states may lead to an underestimate of the importance of those states in the spread of EEEV from Florida, and we are unable to examine the introduction or transmission dynamics in any other region of the country. After Florida, the Northeast has the most reported human and horse cases in the US. It therefore has an outsized importance for understanding the dynamics of EEEV. Obtaining data from other EEEV outbreaks (e.g. Michigan) would provide another opportunity to examine the outbreak dynamics. The sequencing of EEEV samples prior to 2003, a wider geographical range of samples in all time periods, and from Florida after 2014 would greatly add to the reliability and resolution of any phylodynamic study of EEEV.

While some characteristics of the EEEV outbreak in 2019, such as human case traits and climate factors, were within expected ranges, it is clear that many mosquito factors were unusually high (**Fig. 5**). For example, both Massachusetts and Connecticut had a very high abundance of *Cs. melanura* and the vector index for the latter state was the highest ever recorded. While EEEV has a complex ecology, by using data from detailed mosquito surveillance programs, we were able to find strong connections between mosquito infection rate, abundance (connected to climate factors), and human and horse cases. We then developed and applied a negative binomial regression model to utilize early season values of mosquito-specific predictors (most notably the month of first detection and vector index) to provide early estimates of overall case counts for that year. This important development will provide departments of health with estimates to help direct control strategies, and enable more effective communication of risk to the public.

Our models and estimates have several limitations. First, our negative binomial regression model to predict cases is likely more informative for Massachusetts than Connecticut, as there are more cases in Massachusetts to inform it. As such, the Connecticut predictions are heavily influenced by Massachusetts case counts, and given the highly localized nature of EEEV to forested wetland habitats, the model may not accurately represent conditions in Connecticut. In addition, our model was restricted to a monthly time scale to match monthly case data. Given the short duration of the EEEV transmission season, certain dynamics may be missed on the weekly or even daily timescale. There was not enough case data within each month to explore impacts of detection on a month by month basis, making the month detection piece of the analysis less specific. While vector index is a direct risk estimate of EEEV cases, index P had a greater effect estimate in our model. This could be due to our model not fully accounting for the inherent seasonality of EEEV transmission that overlaps with high values of index P or that the true value of index P, which is modeled with a certain degree of uncertainty, lies a significant distance outside the determined estimate. Due to lack of information on EEEV in *Cs. melanura*, index P is estimated using the transmission probability, bird incubation period, bird infectious period and extrinsic incubation period derived from previous West Nile virus research of *Culex pipiens* species (Lourenço et al., 2020). This lack of data for EEEV priors may be confounding the predictive power of this parameter. Finally, veterinary cases are reported voluntarily and the true case counts include interventions undertaken in those years. These interventions include a highly recommended horse vaccination and mosquito spraying: the former will cloud the association between mosquito factors and case counts as exposures may not convert to cases. Both lead to an underestimate of true, unaltered case counts as the model is fitted to partially controlled epidemics.

Despite these limitations, on a short timescale, we have some predictive ability of human and horse cases of EEEV (**Fig. 5**). When considering longer timescales, we still cannot use climate-informed models, like index P, for annual predictions of outbreaks. This is because, while climatic factors are vital for the high abundance of *Cs. melanura* required for an intense EEEV year, they cannot predict when the virus will be present. Index P therefore may be a more effective way to predict EEEV case counts in Florida where the virus is continuously maintained (Bigler et al., 1976), but should be used with caution in scenarios where the virus must be introduced.

To provide longer-term predictions (i.e. on a time scale of years), we must therefore understand what drives introductions of EEEV from Florida into the Northeast. The missing piece here is large-scale studies on bird immunity, as waves of infection in the Northeastern bird population are likely driven by the renewal of the susceptible population, potentially through birth (Armstrong & Andreadis, 2022; Elias et al., 2017). Theoretically, the dynamics could be similar to those of Middle Eastern Respiratory Syndrome (MERS) (Dudas et al., 2018), wherein cycles of infection due to buildup of susceptibles in the reservoir population leads to spillover events into other species. Certainly, we found that EEEV appears to have waves of introductions into the Northeast which co-circulate before going extinct shortly afterwards, which would be expected from local depletion of susceptible birds.

In understanding EEEV outbreaks in humans and horses, we must look to a combination of dynamics of other arboviruses like West Nile and dengue in terms of the mosquito populations, as well as viruses that mostly exist in reservoir populations and spillover into humans, such as MERS. The complex interplay of these factors make long-term prediction with our current data sources difficult, as we do not have enough information on bird immunity and its interaction with EEEV transmission and spillover. We do know, however, that a large mosquito population, enabled by warm and wet conditions, is necessary, and an increase in years with warm and wet summers and mild winters may increase the frequency of outbreaks. Therefore, while the interactions in EEEV with climatic factors are more complex than with some other arboviruses, climate change may still represent an increase in risk as more years will be permissive for outbreaks in mammals. In any case, all of these results rest on a timely and robust mosquito surveillance program, as currently exists in New York, Massachusetts, and Connecticut. Widespread and consistent trapping and rapid analysis provides the data required to calculate vector index, which is the strongest correlate of human and horse cases later in the season. It is imperative that programs like this, also of use for other mosquito-borne viruses such as West Nile virus, continue to be funded and expanded, even as competition for public health funding increases.

## Materials and Methods

### Ethics

No human samples or direct clinical data were used in this study. This study was determined to be Not Human Research by the Yale University Human Research Protection Program Institutional Review Board. All EEEV case data were aggregated and available from public surveillance databases as described below. Sequencing of remnant veterinarian samples by the Wadsworth Center were done following institutional protocols.

### Case data and availability

EEE, caused by EEEV, is a notifiable human disease, therefore human case data is routinely reported to the federal government. Horse data reporting is voluntary and so is likely an underestimate. Data can be accessed on request from ArboNET (https://www.cdc.gov/mosquitoes/mosquito-control/professionals/ArboNET.html).

Base layers for all map figures were taken from the Global Administrative Database (gadm.org).

### Mosquito surveillance

In New York State (NYS), mosquito surveillance was carried out in 13-43 counties throughout the state including an EEEV endemic area of Central NYS (Onondaga, Oswego, Oneida Counties), annually from May-October, as previously described (Oliver et al., 2018). Trapping was completed using a combination of Centers for Disease Control and Prevention (CDC) light traps, gravid traps and diurnal resting boxes. Resting boxes were primarily used in EEEV endemic areas to collect *Cs. melanura*. Mosquito specimens were sorted by species and pooled for testing. Pools of 10-60 mosquitoes were shipped on dry ice to the NYS Arbovirus Laboratory for EEEV testing by molecular and cellular methods. Specifically, pools containing a zinc-plated steel bead and 1ml mosquito diluent (20% heat-inactivated fetal bovine serum (FBS) in Dulbecco’s phosphate-buffered saline plus 50 μg/mL penicillin/streptomycin, 50 μg/mL gentamicin, and 2.5 μg/mL Fungizone) were homogenized using a mixer mill for 30 sec at 24Hz and centrifuged for 5 min at 6000 rcf. Quantitative reverse transcriptase polymerase chain reactions (qRT-PCR) were performed using two distinct primer and probe sets, following RNA extraction and purification, as previously described (Zink et al., 2013). In addition, 100 μL of supernatant from *Cs. melanura* pools were inoculated on Vero cell culture and monitored for cytopathic effect. EEEV isolates obtained from a single round of amplification were used for sequencing.

In Massachusetts, mosquito surveillance was conducted from mid-May through to mid-October, as previously described (Kinsella et al., 2020; Molaei et al., 2013). Trapping was performed by the Massachusetts Department of Public Health (MDPH) in collaboration with 10 local mosquito control projects (MCP) at semi-variable frequencies visiting non-fixed trap sites spread across all 14 counties. Site visitation frequency increased with high volume collections of vector species and narrowed to weekly-biweekly over time in correlation with increased site-specific target mosquito abundance. Targeted sites visitation frequency increased to weekly at minimum when EEEV activity was detected and persisted through the duration of the seasonal surveillance period. Weekly collections were performed at 10 fixed collection sites in Bristol and Plymouth counties known to be historically active EEEV sentinel sites. These site collections increased to twice a week after initial EEEV detection.

Mosquito collection methods varied depending on MCP, however nearly all successful *Cs. melanura* collections were performed using primarily CDC-Miniature Light traps with a CO_2_ source (either dry ice or regulated tank flow ranging from 250-500cc), gravid traps baited with an infusion of lactalbumin-yeast-hay with oak leaves, or resting boxes placed primarily in locales with both deciduous and evergreen freshwater forested swamps. Light traps and gravid traps were placed in the early morning-late afternoon and retrieved 24 hours later, and resting boxes were visited once weekly. Mosquito trap canisters collected from the field were transported to the laboratory in an igloo cooler lined with dry ice, freeze-killed in an ultra-low -80°C freezer, identified by species using a dichotomous key to characterize morphological differences with a stereoscope, and pooled in sample vials of 5-50 female mosquitoes. Sample pools were grouped by species/trap site/date of collection before being submitted to the MDPH Molecular Diagnostics lab for arbovirus testing.

In Connecticut, mosquito trapping and arbovirus surveillance was conducted from the beginning of June through the end of October at 91 fixed collection sites, distributed among all 8 counties. Trapping locations, where *Cs. melanura* were likely to be collected or where there was historical record of EEEV activity, were established in sparsely populated rural settings that included permanent fresh-water swamps (red maple/white cedar) and bogs, coastal salt marshes, horse stables, and swamp-forest border environs. Additional trap sites are located in more densely populated urban or suburban locales, including parks, greenways, golf courses, undeveloped wood lots, sewage treatment plants, dumping stations, and temporary wetlands associated with waterways.

Mosquito trapping was conducted with CO_2_ (dry ice)-baited CDC miniature light traps equipped with aluminum domes or gravid mosquito traps baited with a lactalbumin-yeast-hay infusion. Traps were placed in the field in the afternoon, operated overnight, and retrieved the following morning. Trapping frequency was minimally made once every ten days at each trap site over the course of the entire season. Mosquito trapping frequency was increased at EEEV-positive sites to twice per week after the virus was isolated from that site. Adult mosquitoes were transported alive to the laboratory each morning in an ice chest lined with cool packs. Mosquitoes were immobilized with dry ice and transferred to chill tables where they were identified to species with the aid of a stereo microscope (90X) based on morphological characters. Female mosquitoes were pooled in groups of 50 or fewer by species, collection date, trap type, and collection site and stored at -80°C until processed for virus isolation. Processed mosquito pools were inoculated into Vero cell cultures and screened for cytopathic effect (CPE) as previously described (Armstrong et al., 2011). CPE positive virus cultures and the original mosquito pool were then tested for EEEV by TaqMan RT-PCR assay (Armstrong et al., 2012).

### Maximum-likelihood estimation of infection rate, relative abundance, and vector index

The maximum-likelihood estimation (MLE) of the infection rate is a pooled infection rate of positive EEEV pools, and was estimated using the CDC R software package PooledInfRate (https://github.com/CDCgov/PooledInfRate). This estimation procedure takes into account that there may be different numbers of positive mosquitoes in each positive pool, and so estimates the likely number of positive mosquitoes in each pool based on the overall number of positive pools. The relative abundance was calculated as the average number of mosquitoes captured per trap per night. Vector index is these two metrics multiplied together (Fauver et al., 2016; Nasci et al., n.d.).

### RNA isolation and virus sequencing

RNA was extracted on the MagMax-96 Express robot (Applied Biosystems, Foster City, CA) with the Magmax Viral isolation kit (ThermoFisher Scientific, Waltham, MA), according to manufacturer’s specifications. 50 μL of sample was added to 130 μL of lysis buffer containing 20 μL of RNA binding beads diluted 1:1 with wash buffer. RNA was eluted in 90 μL of elution buffer. Primer pairs, ATAGGGTACGGTGTAGAGGCAACC, TGGTCCGGCATCCCCTTTCTTTAC, and CGTTAACGGAGGGGCACTGAAT, GCGTAGATGCCGGTAGATAACAAC, and AAAGCGCACCTCGTCAAGCATTCT, GCGGTGAGTCTTATCGGGTTTGTC, and CGAAACGGAATTGCAATGTCACTC, CTGATCATAGGCTCGGCTGTCGTA, and CCAAAAGGGGGTTACAGTCAAA, TCGGTGTCGCAGAAGCAGTAGG, and CAAAAGTGCCGTCTCCAGTAGTGA, GAAATATTAAAAACAAAATAAAAACATAAAA, were used to generate 6 overlapping fragments of approximately 2.5 kb each using one-step superscript III RT–PCR with platinum Taq (Life Technologies, Carlsbad, CA). Each reaction utilized 5 μL of RNA, 1 μL of enzyme, and a 0.4 μM final concentration of primer pairs in a total reaction volume of 25 μL. Thermocycler amplification was completed using the following conditions: 55°C for 30 min; 94°C for 2 min; 40 cycles of 94°C for 15 sec, 55°C for 30 sec, 68°C for 3.5 min; and a final extension of 68 °C for 10 min (Simpliamp by Applied Biosystems, Waltham, MA). Two uL of amplicons were visualized on a 1% agarose gel to confirm size and quality, and subsequently purified using Zymo DNA Clean and Concentrate (Zymo Research, Irvine, CA). Amplicons from individual isolates were pooled and sent to the Wadsworth Center Advanced Genomic Technologies Core for library preparation and indexing using the Nextera XT kit (Illumina, San Diego, CA) according to manufacturer’s protocols.

Sequencing was performed on the Illumina MiSeq platform (San Diego, CA). Paired-end reads were assembled to a 2014 Connecticut isolate from *Cs. melanura* (KX029260) deploying Geneious Prime’s reference mapping tool with high sensitivity and free end gaps using 10 iterations of fine tuning and trimming paired read overhangs. Mean coverage/base ranged from 703-2132x. Resultant consensus sequences were used for downstream analyses. We generated complete genome consensus sequences for all 80 sequenced isolates.

### Nucleotide alignments and phylogenetic analysis

In total, 80 new samples of EEEV were sequenced from 2015-2019. These were deposited in Genbank with accession numbers OQ511733-OQ511812. 451 additional whole genome sequences from prior to this study were downloaded from Genbank. Those not from the US and without location data were removed, and the remaining combined with the new sequences to give a dataset of 523 sequences.

We aligned the sequences using Mafft version 1.3.7 (Katoh & Standley, 2013), and removed the non-coding regions at either end of the genome, giving a final multiple sequence alignment with a length of 11,277 bases. A maximum-likelihood phylogenetic tree was generated using IQ-TREE 2.1.4 (Minh et al., 2020) and temporal signal was assessed in TempEst (Rambaut et al., 2016). A single molecular clock outlier (ID: AY722102) was removed, as described in (Hill & Baele, 2019).

We estimated introductions into the Northeast (defined as New York, Connecticut, Massachusetts, New Hampshire, Vermont, Rhode Island and Maine) by conducting a discrete trait phylogeographic analysis (DTA) at the state level in BEAST 1.10 (Suchard et al., 2018), with an asymmetric CTMC model. We also used a non-parametric skygrid coalescent model (Gill et al., 2012) estimated using Hamiltonian Monte Carlo sampling (Baele et al., 2020), an HKY substitution model (Hasegawa et al., 1985), and a strict clock model (Ferreira & Suchard, 2008). We used tip-date sampling for those sequences without exact sampling dates, with a starting date given as 0.6 of the way through the appropriate year (i.e. August) with a standard deviation of three months. We estimate Markov jump histories for the full posterior to obtain estimates of location transitions between states in the DTA, and summarize them using TaxaMarkovJumpHistoryAnalyzer (Lemey et al., 2020). We performed two independent replicates of this analysis, with each chain running for 100 million iterations, removing 10% for burn-in. Convergence and mixing were assessed in Tracer 1.7 (Rambaut et al., 2018).

An introduction node was considered to be the first node which was inferred to be in one of the Northeastern states and downstream tips were counted as part of the cluster. One introduction left Massachusetts and returned to Florida. As this is unlikely given bird migration patterns, and the confidence in the location of the node was low (52% Massachusetts, 46% Florida), we instead used the child node which was found in Massachusetts, thereby excluding the Florida sequence from this cluster.

As there are no sequences from Florida (which was estimated to be the backbone of the phylogeny) after 2014, and relatively few before, the DTA will underestimate the number of introductions in total as they will not be broken up by Florida sequences. The date of the introduction node (the first node in the cluster inferred to be in the Northeast) will therefore also be too far in the past, as the lack of Florida sequences in the cluster means that the location of the node is inferred incorrectly. Therefore, we set out to split up some introductions which contained long branches and likely represented unsampled Floridian diversity. We identified all of the clusters which contained tips only from the Northeast with more than three sequences and more than 50% of the sequences sampled in 2014 or earlier (i.e. when the last Floridian sequence was sampled); and calculated their average branch length (1.28 years). We then traversed the tree, and for any branches where both the parent and child node were inferred to be in the Northeast, but were more than twice the standard deviation (1.52 years) above the average branch length (threshold = 4.33 years), we assigned the introduction node as the child of the pair instead of the parent. This therefore moved the introduction node closer to the present, and sometimes excluded Northeastern sequences on the sister branch, making separate introductions. The final number of introductions into the Northeast before this procedure was 42 prior to 2014 and 19 after, and 49 and 26 respectively, adding 14 more introductions in total.

To test for nucleotide substitutions common to 2019 sequences, we compared the consensus sequences to the reference sequence used in (Yu et al., 2015), which has Genbank accession ID KJ469556.

### Effective population size using a skygrid model

To identify any possible associations between effective population size and case counts, we applied the latter as covariates to the estimation of population size using the skygrid model (Gill et al., 2016). We began by comparing 12 possible covariates (all human and animal cases, just human cases, just horse cases, and all cases in Massachusetts, Connecticut, and New York with 0, 1 and 2 year lags) to effective population estimated sizes using a skygrid model with no covariates (Gill et al., 2012). On the basis of this preliminary analysis, we ran formal analyses with all cases, human cases, and horse cases and the relevant lags. We also only used sequence and case data from 2003 onwards (n=423), when the surveillance program began, in order to eliminate some noise from the data. BEAST runs were set up as above with tip-date sampling, HKY substitution models and a non-parametric skygrid coalescent model. Grid points were externally set to correspond to the start of each year.

Case counts for years between the inferred time of origin of the tree and 2003 were estimated independently using a normal prior, whose mean is the recorded case count. The standard deviation was calculated such that all national cases and horse case covariates were allowed to have ±5 cases in the 95% confidence interval; and human covariates were allowed to have ±1 cases in the same interval, as the recording of the latter is more precise. This was also undertaken for 2021 data for both analyses with the 2 year lag covariates, as finalized data has not been released by ArboNET at time of writing.

Starting values for unobserved years were taken from news reports and other publicly available sources. Most of the researched outbreaks were reported in (Corrin et al., 2021), and data were supplemented by going to each of the references given in that paper. Data for outbreaks in Michigan were supplemented by information in (Stobierski et al., 2022), and Massachusetts data were supplemented using (Feemster, 1938; Grady et al., 1978; Massachusetts Department of Public Health, 2022). Data for all states were supplemented using CDC Morbidity and Mortality Weekly reports in relevant years. Horse cases used are both confirmed and suspected, due to a lack of equine testing, especially in the earlier epidemics.

### Human outdoorness measurements

How much time people spent indoors or outdoors was taken from (Susswein et al., 2022), who used anonymized GPS data from mobile phones. This data was only available for 2018 and 2019. We took the average indoor/outdoorness for each county which had a case in 2019, or had ever had a case and performed a dependent t test for paired samples to compare the metric between years.

### Bird abundance

Bird banding data to calculate abundance and proportion were obtained from the North American Bird Banding Program dataset (Celis-Murillo et al., 2022). All ages of birds were used. Bird species were selected using data from (Molaei et al., 2016), who calculated the relative feeding preference of *Cs. melanura* in Connecticut using a blood meal analysis and normalizing it by abundance of the bird species in question. High-preference birds in this analysis were the 8 species identified in this paper, and low-preference birds were randomly selected from their list of birds for which *Cs. melanura* appeared to have no preference. We conducted Poisson regressions for each combination for the abundance or proportion of a bird species in a given year against the human and horse cases or mosquito infection rate (both defined above).

### Mosquito transmission suitability model - index P

Index P is an estimate of the transmission potential by a single female mosquito using Bayesian methods and mosquito-virus specific priors (**Table S3**), including transmission probability as a function of temperature and relative humidity. Unlike the basic reproduction number, R_0_, index P does not need to be greater than one to cause sustained transmission, as index P is multiplied by the ratio of human to mosquitoes to derive R_0_. We estimated the variable index P using the Mosquito Virus Suitability Estimator (MVSE), an R package to download the functions for modeling the suitability of a given environment for mosquito-borne virus transmission (Obolski et al., 2019). Temperature and humidity data for each state were calculated by taking the average temperature for each day in the center of each county in the state. These county level data were then averaged to obtain a single temperature estimate for the entire state. All weather data were provided by visualcrossing.com. Index P was calculated from April to October, in line with the transmission season for EEEV. All negative temperatures were set to zero in the model to avoid erroneous results, as at zero or below, mosquitoes are in a hibernation state and so there is no mosquito activity that varies by temperature.

We used a 1 month lag time from index P to cases to account for delays caused by requiring transmission into humans and horses, followed by incubation times and time to diagnosis. Therefore the relevant mosquito activity will be before human and horse cases are reported.

While index P is seen as an important covariate in the negative binomial regression model, it is difficult to tease out the seasonality of the mosquito season with cases. Due to the sporadic nature of EEEV in the Northeast, index P is a necessary but not sufficient parameter to consider. To test whether temperature could be used as a proxy, an additional model was tested by replacing index P with temperature which resulted in a slightly higher Akaike information criterion (AIC) score (231.4 vs 232.3) and wider confidence interval. Given the widespread use of modeled mosquito viability parameters, we opted to keep index P. However, future research and surveillance programs may wish to utilize temperature, an easily obtained parameter with little uncertainty in its estimate.

### Regression model fit to cases

We restricted the case data (described above) to Connecticut and Massachusetts for the model where we had sufficient mosquito data to calculate the Vector Index. Dispersion was detected with a dispersion parameter alpha = 0.18 (p-value = 0.019) using the dispersion test from the AER package in R (Cameron & Trivedi, 1990). This led us to fit a negative binomial regression model (estimated using ML) to predict human_equine with vector_index (formula: human_equine ∼ vector_index + month_detected_july + indexP_lag1 + st_grp + year_index + month_f).

Given the vast differences in cases between Massachusetts and Connecticut we attempted to run stratified models of each state separately. However the model failed to converge for Connecticut, likely due to low availability of cases. Thus a model combining both was used. Months were included as categorical variables with the months May through July as reference groups. These months were combined due to May and June having no cases, making parameter estimates fail to converge when separate. The parameter ‘first month detected July’ was a binary variable determined by identifying the month when the first non-zero value for the vector index occurred for each year. All continuous data were normalized so interpretation of estimates are for a 1 standard deviation increase in the term.

Effect modification of vector index by month of detection was explored in addition to the original model without effect modification. AIC scores of the model with effect modification and without were nearly identical (231.4 vs 231.2 respectively) and an ANOVA test was conducted between the two models and found to have no difference (p-value = 0.14). While no difference was identified, this may be due in part to the small sample size of years where the month detected was after July.

In addition to effect modification, given some of the complexity of index P, we explored the terms temperature 1-month lag and mosquito abundance 1-month lag in replacement of index P. Utilizing the two point rule of thumb for AIC, the abundance model performed worse (231.4 vs 233.2) and its estimate was not significant (CI 95%: 0.57-1.62). The temperature performed slightly worse but similar to the index P model (AIC 231.4 vs 232.3) but had a larger standard error (CI 95%: 1.15-9.31). Given the desire to explore the utility of index P and its higher performance we opted to focus on this model. However, for simplicity sake future work may utilize temperature for its ease of use and similar performance (Figure S6 and Table S4).

Utilizing the DHARMa version 0.4.6 package in R (https://CRAN.R-project.org/package=DHARMa), we simulated the residuals 1,000 times to test for heteroskedasticity, zero inflation, and autocorrelation. Upon visual inspection no heteroskedasticity of the residuals was detected. The simulated ratio of expected to actual zeros was 1.01 (p-value = 0.776) The Durbin-Watson test for autocorrelation was conducted on a subset of the Massachusetts data to avoid duplicate time indexes. In addition to subsetting the data, index P with and without a 1-month lag was tested per the suggestion of the Durbin-Watson test to avoid lagging covariates. Autocorrelation was borderline but insignificant without the index P lag (DW = 1.59, p-value = 0.051).

## Data Availability

Human and horse case data are available on request to ArboNET https://wwwn.cdc.gov/arbonet/maps/ADB_Diseases_Map/index.html.
Genomic data is available online at Genbank, accession numbers OQ511733-OQ511812.
Mosquito data is available upon reasonable request to authors.
Other data is available at https://github.com/grubaughlab/eeev-genomics

## Acknowledgements

We thank the seasonal mosquito surveillance programs from Connecticut, Massachusetts, and each county in New York, Wadsworth Center Advanced Genomic Technologies Core for assistance with sequencing, D. Weinberger for help with the statistical model, J. Howard for his thoughts, and P. Jack and S. Taylor for providing comments on an early draft. Research reported in this publication was supported by the National Institute Of Allergy And Infectious Diseases of the National Institutes of Health under Award Number DP2AI176740 (NDG), CTSA Grant Number UL1 TR001863 from the National Center for Advancing Translational Science (NCATS), a component of the National Institutes of Health (CBFV), the Cooperative Agreement Number U01CK000509 funded by the Centers for Disease Control and Northeast Center of Excellence (PMA, ATC), the KU Leuven Internal Funds Grant No. C14/18/094 (GB), and the Research Foundation - Flanders award “Fonds voor Wetenschappelijk Onderzoek - Vlaanderen,” G0E1420N, G098321N (GB). The content is solely the responsibility of the authors and does not necessarily represent the official views of the National Institutes of Health or the Centers for Disease Control and Prevention.

## Data availability

New sequences were deposited in NCBI Genbank under accession numbers OQ511733-OQ511812. Case data from the literature is available at https://github.com/grubaughlab/eeev-genomics, along with XMLs for the BEAST analyses. ArboNET case data is available on request from the CDC https://wwwn.cdc.gov/arbonet/maps/ADB_Diseases_Map/index.html. Original shape files available from gadm.org.

