## Supplementary Information for "Dynamics of Eastern equine encephalitis virus during the 2019 outbreak in the Northeast United States"

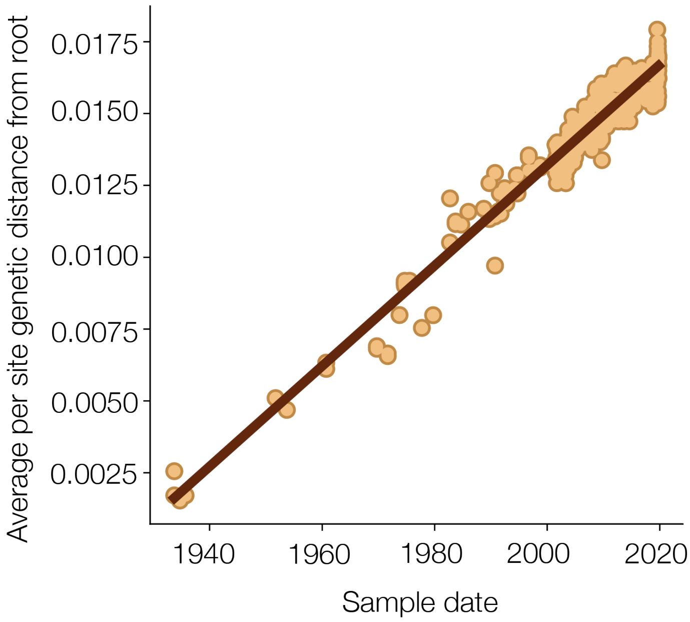


***Figure S1 | Regression of EEEV sequence collection dates against genetic distance from the root of the phylogeny.*** *Each dot represents a tip in the phylogeny, and the gradient of the line gives the rate of evolution of 1.74 x* $10^{-4}$*. The low scatter of the dots around the line indicates a strong temporal signal.*

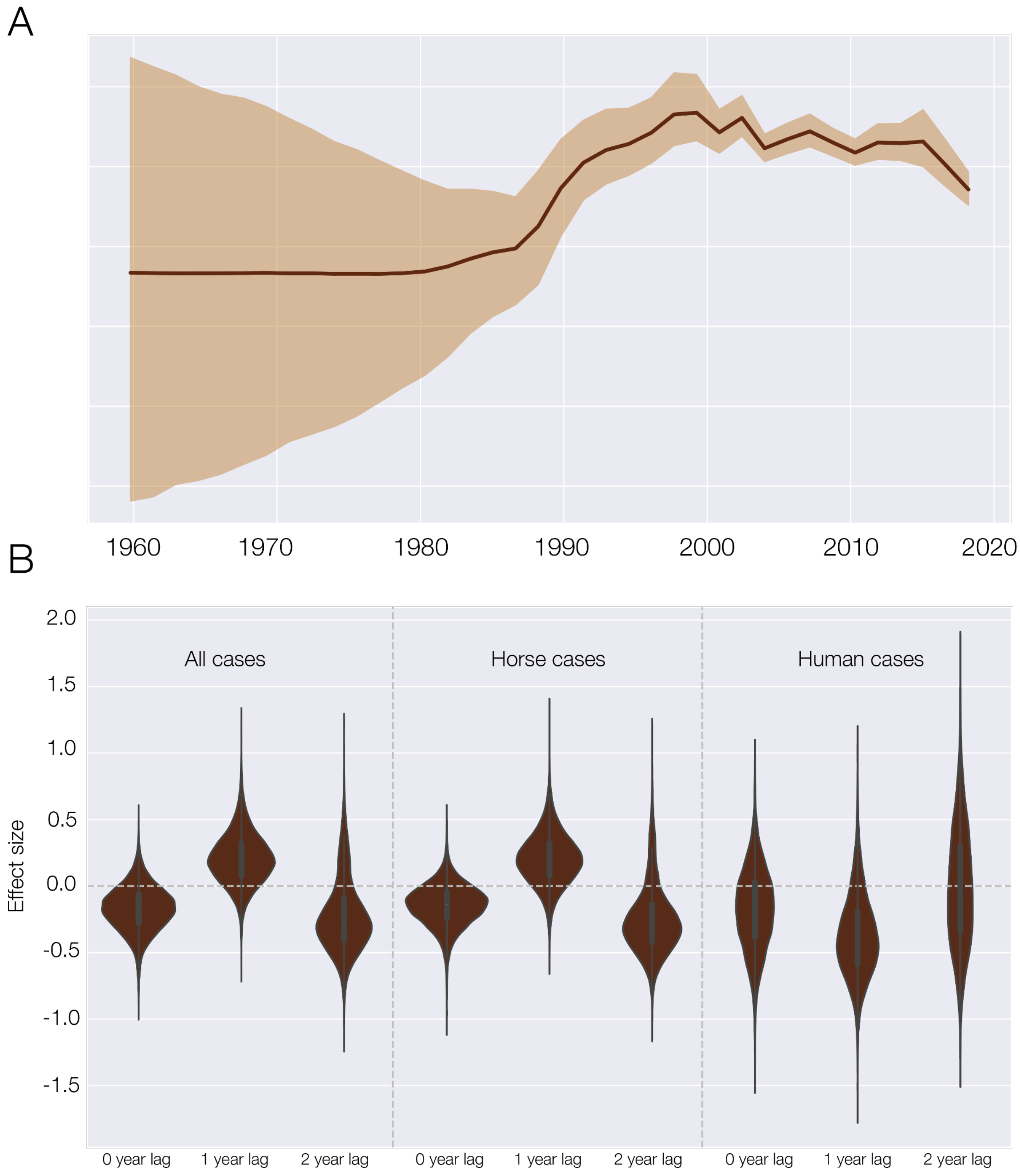


***Figure S2 | Virus effective population size*** *A) Change in viral effective population size over time as estimated by a non-parametric skygrid coalescent model. B) Kernel density estimates of the effect size of various case covariates on the historical change in population size of EEEV.*

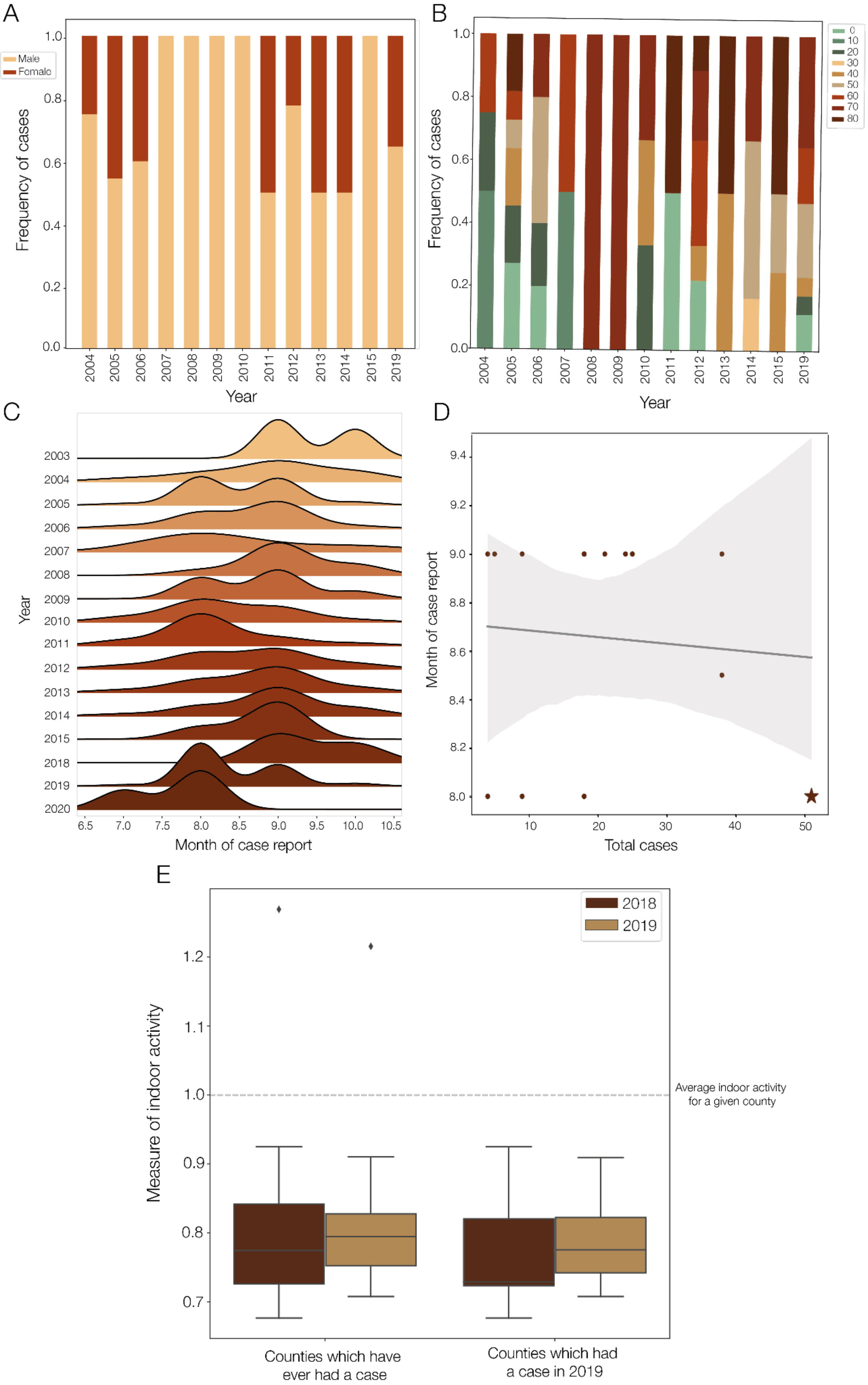
A
Male
Female
Frequency of cases
Year
B
Frequency of cases
Year
C
Year
Month of case report
D
Month of case report
Total cases
E
2018
2019
Measure of indoor activity
Average indoor activity
for a given county
Counties which have
ever had a case
Counties which had
a case in 2019

***Figure S3 | Human case characteristics and behavior.*** *A) Frequency of sex of human cases by year. B) Frequency of human cases in each age category by year. C) Kernels of human and horse case counts over time per year. D) Regression of total cases against median month of symptom onset of human and horse cases; there is no relationship between the two. E) Measure of indoor activity for counties in Massachusetts, Connecticut, and New York which have ever had a case or only had a case in 2019, coloured by year. The dotted line at 1.0 indicates the average indoorness of a county, and values less than 1.0 indicate more indoor activity than average.*

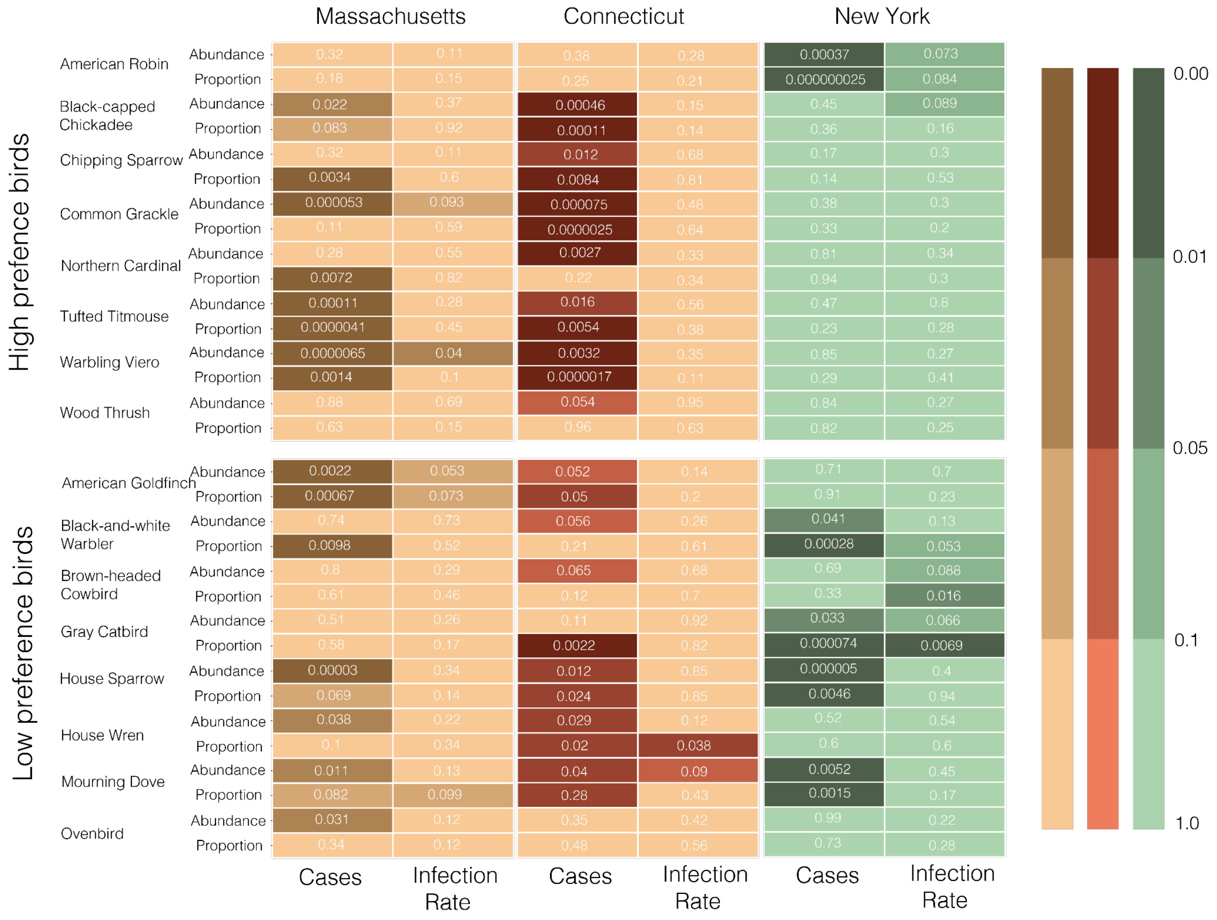


***Figure S4 | Bird abundance and proportion in the Northeast.*** *Heatmap showing p-values of Poisson regressions by state of the proportion and abundance of birds which Cs. melanura has a high or low preference for against the human and horse cases or infection rate. Light colors indicate a lower p-value, and boxes are annotated with the p-value.*

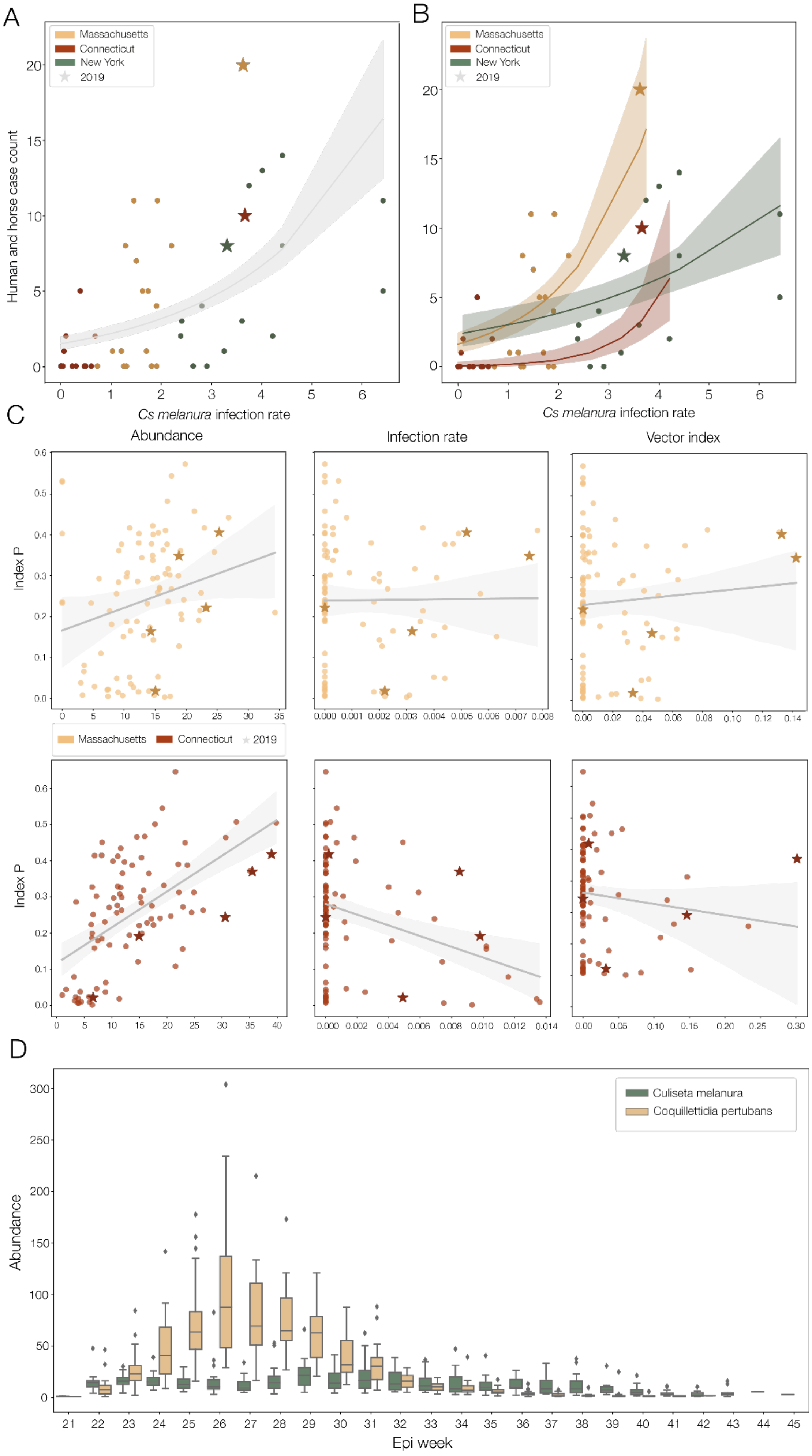
A
B
Massachusetts
Connecticut
New York
2019
Human and horse case count
Cs melanura infection rate
C
Abundance
Infection rate
Vector index
Index P
D
Culiseta melanura
Coquillettidia pertubans
Abundance
Epi week

***Figure S5 | Mosquito infection rate and abundance data compared to index P and case counts.*** *Poisson regression of Cs. melanura infection rate against human and horse case count A) overall and B) by state. In both, data from 2019 are shown as stars. C) Linear regressions of index P against abundance, IR and VI for Connecticut and Massachusetts. Index P and Abundance have the strongest correlation. D) Abundance of Cs. melanura and Cq. perturbans from 2003-2019 across the year from week 21 (approximately mid-May depending on year) to week 45 (approximately the end of October, depending on year).*

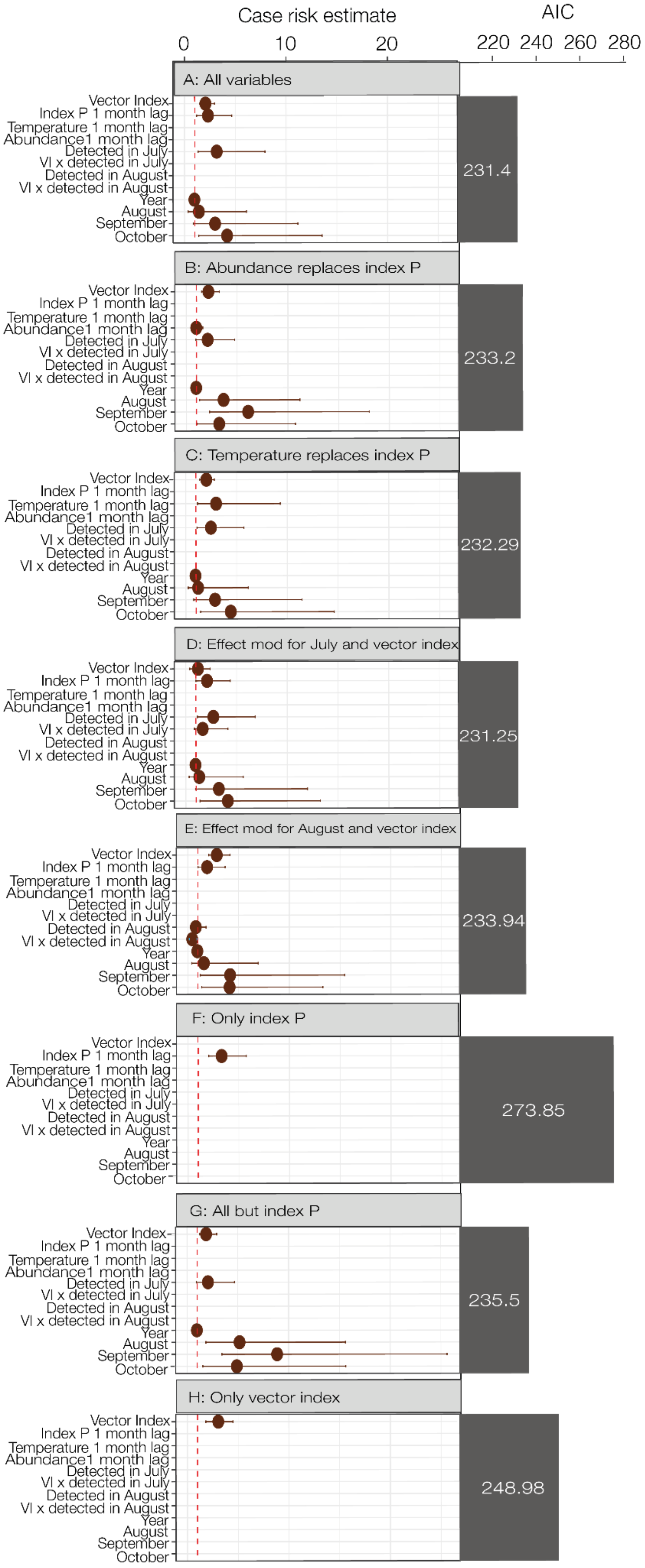

Case risk estimate
0
10
20
AIC
220
240
260
280
A: All variables
231.4
B: Abundance replaces index P
233.2
C: Temperature replaces index P
232.29
D: Effect mod for July and vector index
231.25
E: Effect mod for August and vector index
233.94
F: Only index P
273.85
G: All but index P
235.5
H: Only vector index
248.98
Vector Index
Index P 1 month lag
Temperature 1 month lag
Abundance1 month lag
Detected in July
VI x detected in July
Detected in August
VI x detected in August
Year
August
September
October

***Figure S6 | Results of different negative binomial models to explain cases**. Brief description of each model above the relevant panel. Akaike information criteria (AIC) results shown above. The best fitting models here using this criterion is model D or model A, as they have the lowest AIC value. Results can be found in Table S4. Model A is the same as that shown in Figure 5C and its results are in Table S3. State (i.e. risk of being in Massachusetts compared to Connecticut) is not shown for scale.*

*Table S1 | Metadata for sequenced EEEV isolates*

| Name | Fasta name | Accession | Location | Host | Date |
|---|---|---|---|---|---|
| 14117-19 | 14117-19_S75_2019\|2019-09-23 | OQ511757 | Connecticut: New_Haven | Culiseta melanura | 2019-09-23 |
| 11403-19 | 11403-19_S64_2019\|2019-08-27 | OQ511749 | Connecticut: Middlesex | Culiseta melanura | 2019-08-27 |
| 14149-19 | 14149-19_S76_2019\|2019-09-23 | OQ511752 | Connecticut: Middlesex | Culiseta melanura | 2019-09-23 |
| 10229-19 | 10229-19-19_S57_2019\|2019-08-19 | OQ511779 | Connecticut: Middlesex | Culiseta melanura | 2019-08-19 |
| 14759-19 | 14759-19_S80_2019\|2019-09-30 | OQ511781 | Connecticut: New_London | Culiseta melanura | 2019-09-30 |
| 12004-19 | 12004-19_S68_2019\|2019-08-29 | OQ511780 | Connecticut: New_London | Culiseta melanura | 2019-08-29 |
| 10529-19 | 10529-19_S59_2019\|2019-08-20 | OQ511748 | Connecticut: Middlesex | Coquellitidia perturbans | 2019-08-20 |
| 10084-19 | 10084-19_S56_2019\|2019-08-15 | OQ511745 | Connecticut: Windham | Culiseta melanura | 2019-08-15 |
| 14661-19 | 14661-19_S79_2019\|2019-09-26 | OQ511746 | Connecticut: Windham | Culiseta melanura | 2019-09-26 |
| 14098-17 | 14098-17_S47_2017\|2017-10-11 | OQ511793 | Connecticut: Windham | Culiseta melanura | 2017-10-11 |
| 14229-17 | 14229-17_S49_2017\|2017-10-25 | OQ511754 | Connecticut: Windham | Culiseta melanura | 2017-10-25 |

| | | | | | |
|---|---|---|---|---|---|
| 17667-18 | 17667-18_S50_2018\|2018-09-19 | OQ511794 | Connecticut: Windham | Culiseta melanura | 2018-09-19 |
| 10278-19 | 10278-19_S58_2019\|2019-08-19 | OQ511747 | Connecticut: Middlesex | Culiseta melanura | 2019-08-19 |
| 12670-19 | 12670-19_S71_2019\|2019-09-05 | OQ511759 | Connecticut: New_London | Culiseta melanura | 2019-09-05 |
| 15347-19 | 15347-19_S82_2019\|2019-10-10 | OQ511802 | Connecticut: New_London | Culiseta melanura | 2019-10-10 |
| 19350-18 | 19350-18_S53_2018\|2018-10-09 | OQ511791 | Connecticut: New_London | Culiseta melanura | 2018-10-09 |
| 11716-19 | 11716-19_S66_2019\|2019-08-28 | OQ511751 | Connecticut: New_Haven | Culiseta melanura | 2019-08-28 |
| 15191-19 | 15191-19_S81_2019\|2019-10-07 | OQ511750 | Connecticut: New_Haven | Culiseta melanura | 2019-10-07 |
| 9180-19 | 9180-19_S55_2019\|2019-08-08 | OQ511783 | Connecticut: New_Haven | Culiseta melanura | 2019-08-08 |
| 15435-19 | 15435-19_S83_2019\|2019-10-17 | OQ511755 | Connecticut: Fairfield | Culiseta melanura | 2019-10-17 |
| 10838-19 | 10838-19_S61_2019\|2019-08-21 | OQ511801 | Connecticut: New_London | Culiseta melanura | 2019-08-21 |
| 13289-19 | 13289-19_S72_2019\|2019-09-11 | OQ511805 | Connecticut: New_London | Culiseta melanura | 2019-09-11 |
| 18341-18 | 18341-18_S51_2018\|2018-09-26 | OQ511790 | Connecticut: New_London | Culiseta melanura | 2018-09-26 |
| 14275-19 | 14275-19_S74_2019\| | OQ511782 | Connecticut: New_London | Culiseta melanura | 2019-09-23 |

| | | | | | |
|---|---|---|---|---|---|
| | 2019-09-23 | | | | |
| 11548-19 | 11548-19_S65_2019\|2019-08-27 | OQ511808 | Connecticut: Windham | Culiseta melanura | 2019-08-27 |
| 18694-18 | 18694-18_S52_2018\|2018-10-01 | OQ511792 | Connecticut: Windham | Culiseta melanura | 2018-10-01 |
| 11309-19 | 11309-19_S63_2019\|2019-08-26 | OQ511797 | Connecticut: Fairfield | Culiseta melanura | 2019-08-26 |
| 13866-19 | 13866-19_S73_2019\|2019-09-17 | OQ511734 | Connecticut: Hartford | Culiseta melanura | 2019-09-17 |
| 14311-19 | 14311-19_S78_2019\|2019-09-23 | OQ511758 | Connecticut: Fairfield | Aedes vexans | 2019-09-23 |
| 10579-19 | 10579-19_S60_2019\|2019-08-20 | OQ511800 | Connecticut: New_London | Culiseta melanura | 2019-08-20 |
| 12388-19 | 12388-19_S69_2019\|2019-09-03 | OQ511803 | Connecticut: New_London | Culiseta melanura | 2019-09-03 |
| 19VX2817 | 19VX2817_S42_2019\|2019-08-28 | OQ511778 | Connecticut: Tolland | Equus caballus | 2019-08-28 |
| 8117-19 | 8117-19_S54_2019\|2019-07-31 | OQ511799 | Connecticut: New_London | Culiseta melanura | 2019-07-31 |
| 10898-19 | 10898-19_S62_2019\|2019-08-21 | OQ511806 | Connecticut: New_London | Culiseta melanura | 2019-08-21 |
| 11854-19 | 11854-19_S67_2019\|2019-08-28 | OQ511807 | Connecticut: New_London | Culiseta melanura | 2019-08-28 |
| 12606-19 | 12606-19_S70_2019\|2019-09-04 | OQ511804 | Connecticut: New_London | Culiseta melanura | 2019-09-04 |

| | | | | | |
|---|---|---|---|---|---|
| 11357-16 | 11357-16_S45_2016\|2016-09-12 | OQ511753 | Connecticut: New_London | Culiseta melanura | 2016-09-12 |
| 13966-17 | 13966-17_S46_2017\|2017-10-05 | OQ511789 | Connecticut: New_London | Culiseta melanura | 2017-10-05 |
| CC19-0533 | CC19-0533_S36_2019\|2019-09-10 | OQ511738 | Massachusetts: Barnstable | Culiseta melanura | 2019-09-10 |
| CC19-0548 | CC19-0548_S37_2019\|2019-09-12 | OQ511741 | Massachusetts: Barnstable | Culiseta melanura | 2019-09-12 |
| SL19-1319 | SL19-1319_S32_2019\|2019-08-13 | OQ511739 | Massachusetts: Bristol | Culiseta melanura | 2019-08-13 |
| SL19-0635 | SL19-0635_S29_2019\|2019-07-22 | OQ511743 | Massachusetts: Bristol | Culiseta melanura | 2019-07-22 |
| NM19-0698 | NM19-0698_S39_2019\|2019-09-23 | OQ511786 | Massachusetts: Essex | Culiseta melanura | 2019-09-23 |
| SL19-1569 | SL19-1569_S35_2019\|2019-08-29 | OQ511798 | Massachusetts: Hampden | Culiseta melanura | 2019-08-29 |
| SL19-1219 | SL19-1219_S41_2019\|2019-08-07 | OQ511787 | Massachusetts: Norfolk | Culiseta melanura | 2019-08-07 |
| SL19-1332 | SL19-1332_S31_2019\|2019-08-13 | OQ511740 | Massachusetts: Plymouth | Coquellitidia perturbans | 2019-08-13 |
| SL19-0643 | SL19-0643_S40_2019\|2019-07-22 | OQ511742 | Massachusetts: Plymouth | Culex salinarius | 2019-07-22 |
| SL18-1166 | SL18-1166_S27_2018\|2018-09-17 | OQ511785 | Massachusetts: Plymouth | Culex pipiens-restauns | 2018-09-17 |
| PY19-0166 | PY19-0166_S30_201 | OQ511812 | Massachusetts: Plymouth | Culiseta melanura | 2019-07-23 |

| | | | | | |
|---|---|---|---|---|---|
| | 9\|2019-07-23 | | | | |
| SL18-1204 | SL18-1204_S28_2018\|2018-09-26 | OQ511735 | Massachusetts: Worcester | Culex pipiens-restauns | 2018-09-26 |
| 18VX3174 | 18VX3174_S43_2018\|2018-11-01 | OQ511736 | Massachusetts: Worcester | Equus caballus | 2018-11-01 |
| 18VX2715 | 18VX2715_S44_2018\|2018-09-19 | OQ511733 | Massachusetts: Worcester | Meleagris gallopavo | 2018-09-19 |
| SL19-1516 | SL19-1516_S33_2019\|2019-08-25 | OQ511795 | Massachusetts: Worcester | Coquellitidia perturbans | 2019-08-25 |
| SL19-1584 | SL19-1584_S34_2019\|2019-08-29 | OQ511775 | Massachusetts: Worcester | Culiseta melanura | 2019-08-29 |
| CM19-2280 | CM19-2280_S38_2019\|2019-09-24 | OQ511788 | Massachusetts: Worcester | Culiseta melanura | 2019-09-24 |
| RAB15ANI04939-01-00 | RAB15ANI04939_2015\|2015-09-02 | OQ511768 | New_York: Franklin | Equus caballus | 2015-09-02 |
| ARB15MS260105 | ARB15MS260105_S5_2015\|2015-07-06 | OQ511771 | New_York: Madison | Culiseta melanura | 2015-07-06 |
| ARB15MS260175 | ARB15MS260175_S6_2015\|2015-07-28 | OQ511772 | New_York: Madison | Coquellitidia perturbans | 2015-07-28 |
| ARB15MS260353 | ARB15MS260353_S12_2015\|2015-09-23 | OQ511774 | New_York: Madison | Culiseta melanura | 2015-09-23 |
| ARB17MS260236 | ARB17MS260236_S17_2017\|2017-09-19 | OQ511761 | New_York: Madison | Culiseta melanura | 2017-09-19 |
| ARB15MS330238 | ARB15MS330238_S4_2015\|2015-06-25 | OQ511773 | New_York: Onondaga | Culiseta melanura | 2015-06-25 |

| | | | | | |
|---|---|---|---|---|---|
| ARB15MS330683 | ARB15MS330683_S8_2015\|2015-09-01 | OQ511737 | New_York: Onondaga | Aedes canadensis | 2015-09-01 |
| ARB17MS330746 | ARB17MS330746_S18_2017\|2017-09-26 | OQ511762 | New_York: Onondaga | Culiseta melanura | 2017-09-26 |
| ARB18MS330453 | ARB18MS330453_S21_2018\|2018-08-07 | OQ511766 | New_York: Onondaga | Coquellitidia perturbans | 2018-08-07 |
| ARB19MS330484 | ARB19MS330484_S3_2019\|2019-08-15 | OQ511809 | New_York: Onondaga | Culiseta melanura | 2019-08-15 |
| ARB18MS330394 | ARB18MS330394_S19_2018\|2018-07-31 | OQ511765 | New_York: Onondaga | Culiseta melanura | 2018-07-31 |
| RAB19ANI05450 | RAB19ANI05450_S3_2019\|2019-08-27 | OQ511744 | New_York: Wayne | Equus caballus | 2019-08-27 |
| ARB18MS350318 | ARB18MS350318_S22_2018\|2018-09-20 | OQ511784 | New_York: Orange | Culiseta melanura | 2018-09-20 |
| ARB15MS370406 | ARB15MS370406_S7_2015\|2015-07-27 | OQ511776 | New_York: Oswego | Culiseta melanura | 2015-07-27 |
| 16370365 | 16370365_S13_2016\|2016-08-17 | OQ511760 | New_York: Oswego | Culiseta melanura | 2016-08-17 |
| ARB17MS370250 | ARB17MS370250_S26_2017\|2017-07-26 | OQ511763 | New_York: Oswego | Culiseta melanura | 2017-07-26 |
| ARB18MS370301 | ARB18MS370301_S20_2018\|2018-07-30 | OQ511767 | New_York: Oswego | Culiseta melanura | 2018-07-30 |
| ARB19MS370212 | ARB19MS370212_S23_2019\|2019-07-08 | OQ511810 | New_York: Oswego | Culiseta melanura | 2019-07-08 |
| RAB19ANI04511 | RAB19ANI04511_S25_2019\| | OQ511764 | New_York: Oswego | Equus caballus | 2019-08-13 |

| | | | | | |
|---|---|---|---|---|---|
| | 2019-08-13 | | | | |
| RAB16ANI06320-01-00 | RAB16ANI06320-01-00_S15_2016\|2016-10-21 | OQ511756 | New_York: Seneca | Equus caballus | 2016-10-21 |
| RAB15ANI05176-01-00 | RAB15ANI05176_2015\|2015-09-24 | OQ511769 | New_York: Saint_Lawrence | Equus caballus | 2015-09-24 |
| ARB17MS510915 | ARB17MS510915_S16_2017\|2017-08-16 | OQ511796 | New_York: Suffolk | Culiseta melanura | 2017-08-16 |
| ARB19MS510595 | ARB19MS510595_S24_2019\|2019-07-31 | OQ511770 | New_York: Suffolk | Culiseta melanura | 2019-07-31 |
| RAB16ANI05362-01-00 | RAB16ANI05362-01-00_S14_2016\|2016-08-23 | OQ511811 | New_York: Ulster | Equus caballus | 2016-08-23 |
| RAB15ANI05065-01-00 | RAB15ANI05065-01-00_S10_2015\|2015-09-12 | OQ511777 | New_York: Steuben | Equus caballus | 2015-09-12 |

***Table S2 | Priors for the Mosquito-borne Viral Suitability Estimator adapted from*** *(Lourenço et al., 2020)* ***for index P estimates in Figure 5.***

| **Prior** | **Mean** | **Standard deviation** |
|---|---|---|
| Bird lifespan | 12 | 2 |
| Bird to Mosquito Transmission Probability | 0.5 | 0.01 |
| Bird Infectious Period | 6 | 1 |
| Bird Incubation Period | 1.5 | 1 |
| Mosquito Lifespan | 10 | 2 |
| Mosquito Extrinsic incubation period | 4 | 1 |

| | | |
|---|---|---|
| Mosquito Biting rate | 0.14 | 0.02 |

***Table S3 | Results from the negative binomial regression model in Figure 5.***

| Term | Estimate | Std. Error | Conf. Low | Conf. High |
|---|---|---|---|---|
| Vector Index | 2.069 | 0.16 | 1.501 | 2.956 |
| Month Detected | 3.17 | 0.445 | 1.337 | 7.924 |
| Year | 0.967 | 0.034 | 0.906 | 1.03 |
| August | 1.408 | 0.729 | 0.342 | 6.089 |
| September | 3.003 | 0.665 | 0.871 | 11.158 |
| October | 4.184 | 0.569 | 1.39 | 13.551 |
| Index P 1 month lag | 2.312 | 0.33 | 1.193 | 4.654 |
| Massachusetts | 11.948 | 0.623 | 3.879 | 43.699 |

***Table S4 | Results from the additional negative binomial regression model variations. The letter in the Model column relates to the panel in Figure S6.***

| Model | Term | Estimate | Std.Error | Conf. Low | Conf. High |
|---|---|---|---|---|---|
| B: Abundance replaces index P | Vector Index | 2.176 | 0.179 | 1.507 | 3.261 |
| B: Abundance replaces index P | Detected in July | 2.094 | 0.413 | 0.94 | 4.763 |
| B: Abundance replaces index P | Year | 0.96 | 0.038 | 0.892 | 1.032 |
| B: Abundance replaces index P | August | 3.659 | 0.552 | 1.292 | 11.183 |
| B: Abundance replaces index P | September | 6.078 | 0.532 | 2.277 | 18.024 |
| B: Abundance replaces index P | October | 3.228 | 0.578 | 1.041 | 10.735 |

| | | | | | |
|---|---|---|---|---|---|
| B: Abundance replaces index P | Abundance 1 Month Lag | 0.972 | 0.246 | 0.573 | 1.621 |
| B: Abundance replaces index P | Massachusetts | 14.295 | 0.626 | 4.719 | 52.542 |
| C: Temperature replaces index P | Vector Index | 2.025 | 0.16 | 1.465 | 2.908 |
| C: Temperature replaces index P | Detected in July | 2.473 | 0.419 | 1.103 | 5.722 |
| C: Temperature replaces index P | Year | 0.948 | 0.035 | 0.888 | 1.011 |
| C: Temperature replaces index P | August | 1.207 | 0.814 | 0.24 | 6.153 |
| C: Temperature replaces index P | September | 2.87 | 0.694 | 0.774 | 11.446 |
| C: Temperature replaces index P | October | 4.42 | 0.574 | 1.453 | 14.621 |
| C: Temperature replaces index P | Temperature 1 Month Lag | 2.982 | 0.512 | 1.156 | 9.313 |
| C: Temperature replaces index P | Massachusetts | 14.612 | 0.605 | 4.99 | 51.143 |
| D: Effect modifier for July and Vector index | Vector Index | 1.395 | 0.316 | 0.588 | 2.595 |
| D: Effect modifier for July and Vector index | Detected in July | 2.72 | 0.463 | 1.142 | 6.842 |
| D: Effect modifier for July and Vector index | Year | 0.95 | 0.037 | 0.887 | 1.016 |
| D: Effect modifier for July and Vector index | August | 1.321 | 0.724 | 0.325 | 5.647 |
| D: Effect modifier for July and Vector index | September | 3.249 | 0.657 | 0.951 | 11.979 |
| D: Effect modifier for July and Vector index | October | 4.133 | 0.566 | 1.388 | 13.246 |

| | | | | | |
|---|---|---|---|---|---|
| D: Effect modifier for July and Vector index | Vector index x Detected in July | 1.666 | 0.347 | 0.849 | 4.147 |
| D: Effect modifier for July and Vector index | Index P 1 Month Lag | 2.29 | 0.324 | 1.191 | 4.574 |
| D: Effect modifier for July and Vector index | Massachusetts | 12.203 | 0.606 | 4.044 | 43.403 |
| E: Effect modifier for August and Vector index | Vector Index | 2.855 | 0.159 | 2.065 | 4.16 |
| E: Effect modifier for August and Vector index | Detected in August | 0.786 | 0.427 | 0.331 | 1.795 |
| E: Effect modifier for August and Vector index | Year | 0.932 | 0.035 | 0.872 | 0.994 |
| E: Effect modifier for August and Vector index | August | 1.574 | 0.735 | 0.376 | 6.929 |
| E: Effect modifier for August and Vector index | September | 4.155 | 0.663 | 1.214 | 15.45 |
| E: Effect modifier for August and Vector index | October | 4.106 | 0.575 | 1.364 | 13.298 |
| E: Effect modifier for August and Vector index | Vector index x detected in August | 0.428 | 0.4 | 0.141 | 0.892 |
| E: Effect modifier for August and Vector index | Index P 1 Month Lag | 1.9 | 0.304 | 1.012 | 3.68 |
| E: Effect modifier for August and Vector index | Massachusetts | 18.315 | 0.571 | 6.601 | 62.483 |
| F: Just index P | Index P 1 Month Lag | 3.315 | 0.224 | 2.046 | 5.735 |
| F: Just index P | Massachusetts | 4.896 | 0.426 | 2.141 | 11.639 |

| | | | | | |
|---|---|---|---|---|---|
| G: No index P | Vector Index | 2.264 | 0.164 | 1.621 | 3.305 |
| G: No index P | Detected in July | 2.05 | 0.413 | 0.919 | 4.672 |
| G: No index P | Year | 0.955 | 0.036 | 0.893 | 1.019 |
| G: No index P | August | 5.161 | 0.552 | 1.848 | 15.598 |
| G: No index P | September | 8.859 | 0.523 | 3.421 | 25.611 |
| G: No index P | October | 4.892 | 0.562 | 1.637 | 15.727 |
| G: No index P | Massachusetts | 16.038 | 0.638 | 5.204 | 59.747 |
| H: Only Vector index | Vector Index | 3.044 | 0.153 | 2.114 | 4.786 |
| H: Only Vector index | Massachusetts | 24.063 | 0.629 | 7.915 | 97.748 |
